# Uncovering personalised glucose responses and circadian rhythms from multiple wearable biosensors with Bayesian dynamical modelling

**DOI:** 10.1101/2022.08.20.22278813

**Authors:** Nicholas E. Phillips, Tinh-Hai Collet, Felix Naef

**Affiliations:** Institute of Bioengineering, School of Life Sciences, Ecole Polytechnique Fédérale de Lausanne, Lausanne, Switzerland; Nutrition Unit, Service of Endocrinology, Diabetology, Nutrition and Therapeutic Education, Department of Medicine, Geneva University Hospitals, Geneva, Switzerland; Diabetes Centre, Faculty of Medicine, University of Geneva, Geneva, Switzerland

**Keywords:** Wearable devices, biosensors, glucose levels, circadian rhythms, mathematical modelling, Bayesian inference, time series

## Abstract

**Motivation:** Wearable biosensors measure physiological variables with high temporal resolution over multiple days and are increasingly employed in clinical settings, such as continuous glucose monitoring in diabetes care. Such datasets bring new opportunities and challenges, and patients, clinicians and researchers are today faced with a common challenge: how to best capture and summarise relevant information from multimodal wearable time series? Here, we aim to provide insights into individual blood glucose dynamics and their relationships with food and drink ingestion, time of day, and coupling with other physiological states such as physical and heart activity. To this end, we generate and analyse multiple wearable device data through the lens of a parsimonious mathematical model with interpretable components and parameters. A key innovation of our method is that the models are learnt on a personalised level for each participant within a Bayesian framework, which enables the characterisation of inter-individual heterogeneity in features such as the glucose response time following meals or underlying circadian rhythms. This framework may prove useful in other populations at risk of cardiometabolic diseases.

**Summary:** Wearable biosensors and smartphone applications can measure physiological variables over multiple days in free-living conditions. We measure food and drink ingestion, glucose dynamics, physical activity, heart rate (HR) and heart rate variability (HRV) in 25 healthy participants over 14 days. We develop a Bayesian framework to learn personal parameters that quantify circadian rhythms and physiological responses to external stressors. Modelling the effects of ingestion events on glucose levels reveals that slower glucose decay kinetics elicit larger postprandial glucose spikes, and we uncover a circadian baseline rhythm of glucose with high amplitudes in some individuals. Physical activity and circadian rhythms explain as much as 40-65% of the HR variance, whereas the variance explained for HRV is more heterogeneous across individuals (20-80%). A more complex model incorporating activity, HR and HRV explains up to 15% additional glucose variability, highlighting the relevance of integrating multiple biosensors to better predict glucose dynamics.

## Introduction

Wearable biosensors and smartphone applications are increasingly used to measure multiple physiological variables, including glucose levels, food consumption, and physical and heart activity. In contrast to traditional lab measurements taken at a single time point, the high-resolution wearable time series data records dynamic changes of physiological variables in response to external perturbations and as a function of the time of day. While this wearable data has the potential to provide a dynamic view of health states^1^, a major challenge in both clinical and research settings is how to extract physiologically meaningful information from wearable time series data, and in particular when multiple data modalities are combined.

Glucose regulation is a prime example of a dynamic and complex physiological system, as the body is confronted with irregular inputs (i.e. food intake, especially of carbohydrates) and controlled glucose uptake by organs (e.g. muscles, liver). As such, glycaemic regulation employs a range of homeostatic mechanisms including the glucose-insulin negative feedback loop, whereby insulin secretion by the pancreas is tightly regulated to avoid both low (hypoglycaemic) or high (hyperglycaemic) levels of glucose^2,3^. Understanding glucose regulation is an important health problem, as long-term chronic hyperglycaemia in diabetes can lead to micro- and macrovascular complications^4^, and glucose levels show a non-linear association with vascular diseases even in populations without diabetes^5–7^.

As glucose homeostasis is inherently dynamic and glucose levels fluctuate throughout the day, continuous glucose monitors (CGMs) have gained popularity due to the high temporal resolution. CGMs measure glucose in interstitial fluid continuously for up to 10-14 days with satisfactory clinical accuracy compared to reference capillary blood glucose values^8,9^. Standardised CGM-derived metrics such as the coefficient of variation (CV) and the time-in-range (the fraction of time spent within the desired range of 3.9-10.0 mmol/L, 70-180 mg/dL) have been adopted in clinical practice to assess glycaemic control in diabetes with insulin treatment^10–13^. At a more fine-grained level, CGMs have been combined with smartphone records of ingestion events to predict postprandial (post-meal) glycaemic responses (PPGR), where the PPGR is often defined as the area under the glucose curve for the two hours following a recorded ingestion event^14–16^.

Nonetheless, neither the standardised CGM metrics nor the PPGR approach provide a complete picture of the entire glucose time series and its fluctuations over the 24-hour clock. Physiological processes in humans, including glucose metabolism, follow circadian rhythms^17–20^, and responses to oral glucose tests are more pronounced in the evening than the morning^21^. A pre-breakfast rise in glucose levels, termed the “dawn phenomenon”, has been observed since the early 1980s and is often linked with a concomitant early morning rise of cortisol^22,23^, but the amplitude and phase of circadian rhythms in baseline glucose levels have thus far not been well described at an individual level. Identifying the relative contribution of the circadian rhythms to the glucose time series would be particularly helpful for the interpretation of the 24-hour CGM report, which is often discussed with patients to identify patterns of low and high glucose values, and to guide treatment^24^.

In addition to glucose, other physiological responses are accessible with biosensors such as heart rate (HR, beats per minute) and heart rate variability (HRV), where HRV is typically quantified with metrics such as the root mean square of successive differences (RMSSD) between heart beats^25^. Epidemiological data has linked low HRV with high glucose levels^26,27^ and a reduction in HRV has been shown to predict the development of autonomic neuropathy before symptom onset amongst diabetic patients^28^. The simultaneous measurement of HR and HRV can provide insights into the autonomic nervous system activity^29^, as HR receives inputs from both the sympathetic nervous system (SNS, the “flight or fight” response) and parasympathetic nervous system (PNS, the “rest and digest” response), while HRV-derived RMSSD metric is dominated by the PNS via vagal nerve activity^30^. Both HR and HRV are modulated by physical activity, which can now also be conveniently measured with a triaxial accelerometer.

Regarding the analysis of wearable data, the methodological approaches depend on the scientific questions and applications. A diverse range of glucose models have been proposed over the last decades^31–33^, ranging from minimal models^34^ to more detailed simulators with dozens of parameters^35^ and neural networks^36–38^. Recent efforts have also attempted to utilise additional multimodal wearable signals to either improve glucose forecasting or provide more accessible proxies for glucose without using CGMs^39–41^. Many of these methods are specialised towards short-range forecasting, which is certainly useful in applications like the artificial pancreas^42^. In a different context, researchers and clinicians need new wearable data analysis tools to perform statistical comparisons between individuals and quantify changes in glucose regulation across multiple time points and different disease states, but such approaches to extract personalised summary metrics from the global recordings remain comparatively unexplored.

In this study, we acquired multiple wearable biosensor data to monitor food and drink ingestion, glucose excursions, physical activity, HR and HRV in individuals in free-living conditions. Our aim was to quantify the role of external perturbations (such as ingestion events and physical activity) and baseline circadian rhythms on a personalised level. We develop models to analyse the wearable data with distinct model components that capture the interactions between physiological variables, 24-hour rhythms and random fluctuations. Individual-specific parameters are learnt within a Bayesian framework, which provides parameter uncertainties and enables statistical comparisons between participants. We subdivide the problem of analysing the multiple signals by creating three successive mathematical models that include different subsets of variables. Our three-tiered modelling reveals the high degree of personalisation across a wide range of metrics even within a healthy population, from glucose decay kinetics, circadian rhythms in baseline glucose levels, dependence between HR and HRV and the boost in explained glucose variance from adding physical and heart activity data. While our study assessed cross-sectional differences between healthy individuals, future studies will be able to re-use the framework to describe personalised longitudinal changes over time in response to interventions and to cardiometabolic diseases.

## Results

### Measuring multivariable physiological time series in free-living conditions

To quantify the personalised dynamics of individuals in free-living conditions, we measured ingestion events, glucose levels, physical activity, HR and HRV for 25 participants over a two-week period. Participants (16 males, 9 females) were young (mean age 33.0 ± SD 11.0), had a normal weight (mean BMI 22.7 ± 2.8 kg/m^2^, 1 person with overweight and 1 person with obesity) and a normal blood pressure (systolic 117.6 ± 11.4 mmHg, diastolic 75.3 ± 7.9 mmHg). Participants were asked to record all food and drink consumption and add a manual free text annotation of the content with the smartphone application MyCircadianClock^43^. Each ingestion event was automatically timestamped by the app. The adherence (defined as at least 2 meals separated by at least 5 hours in a given day^44^ was above 83% for all participants (Supplementary Table 1).

We measured glucose levels continuously using the Abbott FreeStyle Libre Pro CGM device, which records interstitial glucose levels every 15 minutes over a two-week period. As the device is blinded, participants were unable to access their glucose data during the study period thus avoiding feedback on their eating behaviour. Five participants wore two sensors (on different arms), with the aim of validating that parameters estimated from the model were consistent between the two sensors (noted ID .A and .B in the Figures). Physical activity, HR and HRV were measured for each participant over the two-week study period using the CamNTech ActiHeart version 5 device, and the physical and heart activity data was also blinded to participants during the study.

### Multi-wearable time series data reveal complex dynamical responses as a function of external inputs and time of day

For initial data exploration, we superposed the recorded days of glucose data based on time of day and found strong inter-individual heterogeneity in the mean 24-hour pattern, with the highest mean glucose level occurring in the morning, afternoon or evening depending on the individual (Figure 1A, all participants shown in Supplementary Figure 1). These unique 24-hour trends could be caused by either food or drink ingestion (i.e. external perturbations) and/or an underlying circadian baseline rhythm in glucose. This motivated the inclusion of both ingestion events and circadian rhythms in the model of glucose dynamics as separate components with learnable parameters.

**Figure 1.**
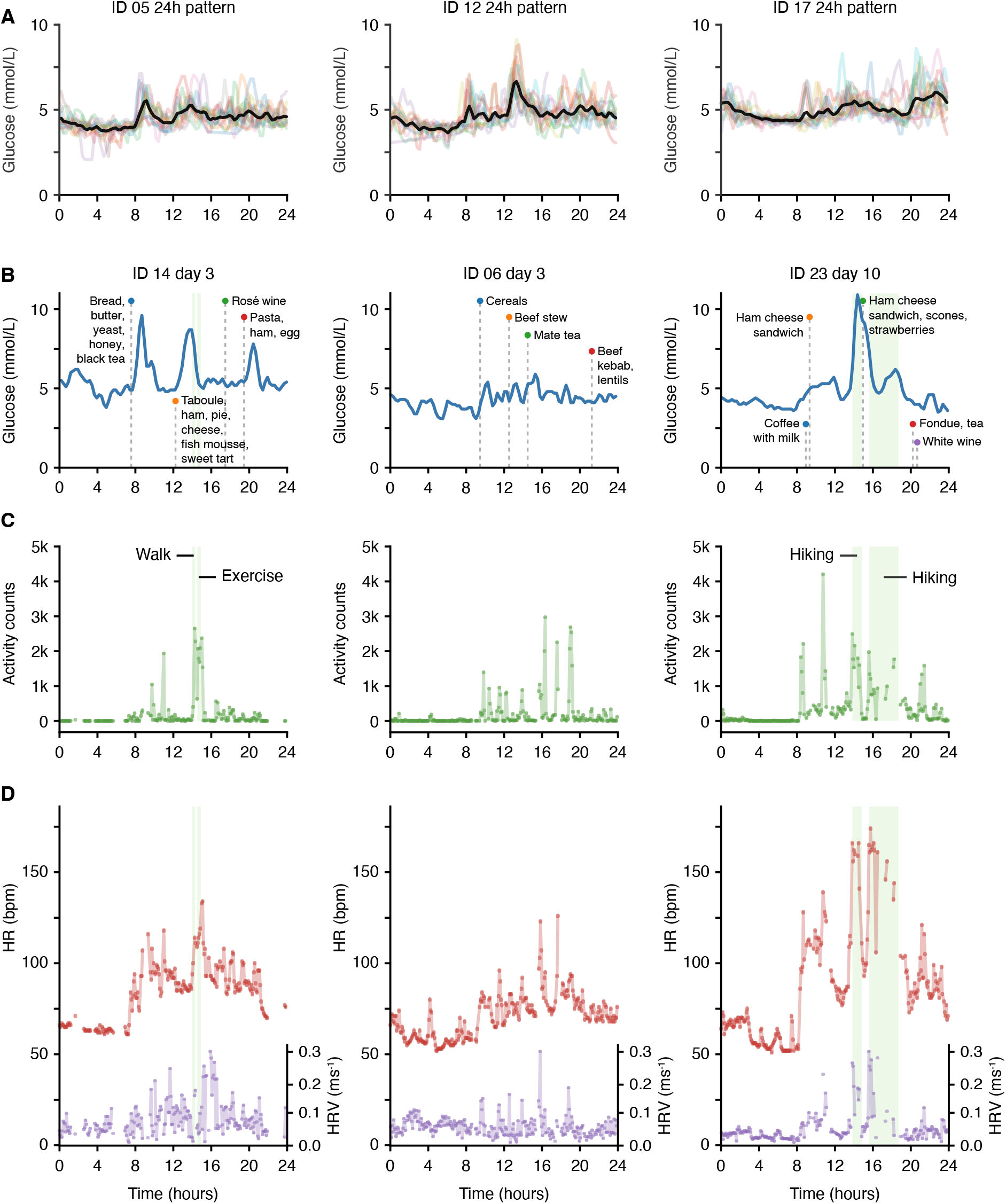
Exploratory analysis of wearable signals: examples of 24-hour trends and responses to external stressors. (A) Continuous glucose monitoring (CGM) data: superposition of all recorded days of data shown on the same 24-hour scale for three different participants (see related Supplementary Figure 1 for all participants). Black: average over all days; coloured lines: data for individual days; time axis: wall clock time. (B) Selected day examples of CGM glucose levels alongside recorded ingestion events for three participants (same individuals shown in panels B-D). Blue: glucose levels; green shade: recorded activity events; time axis: wall clock time; vertical dashed lines: ingestion events. (C) Selected day examples of physical activity measured with the CamNTech Actiheart device. Green: physical activity; green shade: recorded activity events; time axis, clock time. (D) Selected day examples of HR and HRV measured with the CamNTech Actiheart device. Green shade: recorded activity events; purple: heart rate variability (HRV) (quantified with RMSSD^-1^ in ms^-1^); red: heart rate (HR) in beats per min. (bpm); time axis: wall clock time.

Further exploratory analysis of the multiple wearable signals showed rich interactions between the five measured variables (i.e. ingestion, glucose, activity, HR and HRV). As expected, glucose levels often rose following ingestion events, and for some of the individuals, recorded meals seemed to lead to large, predictable peaks in glucose (Figure 1B, ID 14 and 23), while others showed a more complex relationship, with small postprandial glucose spikes that were barely larger than the glucose fluctuations between meals (Figure 1B, ID 06). Based on these observations, and compared to CGM analysis methods that focus exclusively on PPGRs for 2-3 hours^14–16^, our goal is now to dynamically model the entire glucose time series over 2 weeks, including the fluctuating glucose levels occurring overnight or during longer intervals between ingestion events.

Visual inspection of the physical and heart activity data showed that spikes in physical activity typically coincided with an increased HR and HRV (as measured with RMSSD^-1^) (Figure 1C-D). By creating a joint dynamical model of the three signals (physical activity, HR and HRV), we aimed to uncover the inter-individual heterogeneity in the coupling between the multiple signals as well as the underlying circadian rhythms.

Finally, we observed spikes in glucose levels following physical activity for some individuals (Figure 1B-D, ID 23), which could be caused by the release of glucose under the influence of adrenaline/epinephrine or glucagon. However, to establish more firmly whether physical and heart activity signals can explain glucose variation, we developed a dynamical model to mathematically assess the extent to which the total glucose signal across the two-week study period is predictable by the combined meal, physical, HR and HRV data.

### Slow glucose dynamics is associated with large postprandial glucose spikes

The overall data modelling strategy is shown in Figure 2A-C, where we first focus on ingestion events, glucose and circadian rhythms (Model 1, Figure 2A), then the relationships between the physical and heart activity signals (Model 2, Figure 2B) before finally adding interactions from the physical activity and heart signals to the glucose levels (Model 3, Figure 2C). Based on the visual exploration (Figure 1) and physiological knowledge, we first built a minimal dynamical model of glucose levels (Model 1) that included the following four features: i) the ability to produce a continuous postprandial glucose response following an ingestion event, ii) negative feedback (due to the regulating action of insulin, represented as a feedback loop in Figure 2A), iii) a random component that captures the glucose fluctuations between ingestion events and overnight, iv) a circadian baseline rhythm (discussed in the next section).

**Figure 2.**
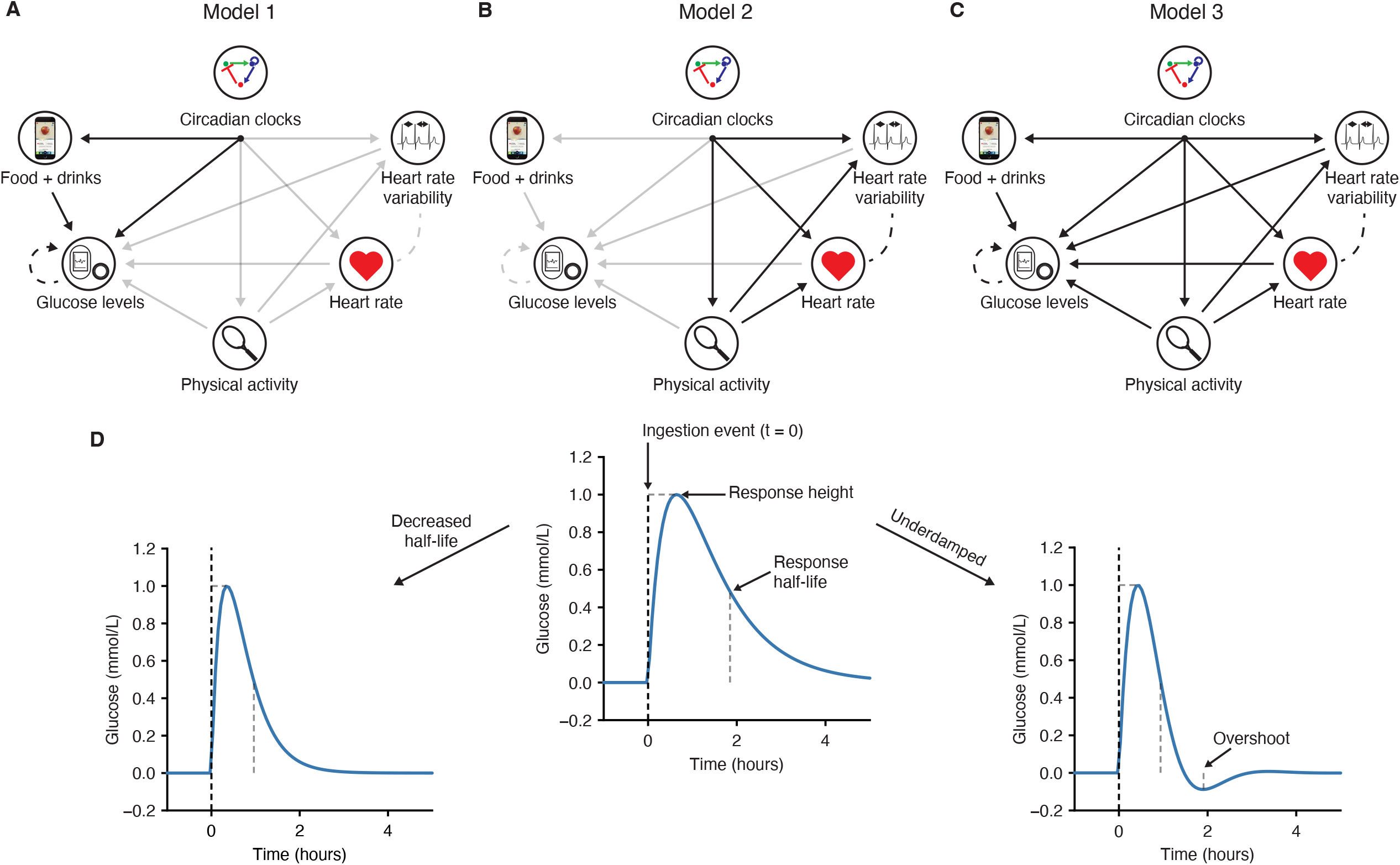
Schematic showing the three different models and parameter interpretation. (A) The glucose and ingestion events interaction (Model 1), (B) the physical and heart activity interaction model (Model 2), (C) the full model (Model 3). Solid arrows represent direct unidirectional influences, while dashed lines represent correlated fluctuations that are not specifically directional. (D) A meal or drink event causes a glucose increase to a specific meal height. The response half-life determines how quickly glucose returns to baseline. Underdamping (defined as a negative damping coefficient) leads to an overshoot below the baseline values.

We modelled the glucose dynamics with a system of stochastic differential equations (SDEs), including negative feedback regulation (see Methods), where ingestion events act to perturb glucose to higher levels. In the deterministic part of the model, after the ingestion causes a glucose increase, glucose levels return to their steady-state values (i.e. reflecting homeostasis). The decay kinetics and precise shape of the response will depend on the parameters of the model (Figure 2D), which are learnt for each participant. Specifically, this individual-specific response to a meal perturbation can be summarised with three parameters, namely a half-life reflecting the time taken for glucose to return to baseline levels, the mean increase in glucose levels caused by meal consumption (referred to as the average meal height) and a damping coefficient specifying whether the response profile is akin to an overdamped (a rapid glucose increase followed by a monotonous slower decay i.e. non-dipping) or an underdamped (leading to a slower initial increase followed by decay and overshoot, i.e. dipping) response. To account for noisy fluctuations in the data, the glucose dynamics is also subjected to random perturbations via a process noise term in the corresponding SDE, meaning that the glucose time series data can show noisy deviations from the idealised meal response.

For each participant, the entire glucose time-series is probabilistically matched (using exact likelihood calculations) to the model using a Gaussian state space model (a.k.a. a Kalman filter), and we infer each of the model parameters using Markov Chain Monte Carlo (MCMC) sampling within a Bayesian framework that yields uncertainty estimates for each parameter (Methods and Supplementary Information).

We first verified model performance by assessing the correlation coefficient between the fitted meal response function and the data (Supplementary Figure 2). The correlation coefficient generally ranged from 0.5 to 0.8 but was particularly low for participant ID 04. Visual inspection of this participant’s raw data showed large glucose spikes following physical activity, which we address at a later stage of the modelling. While all model parameters are shown in Supplementary Figure 2, here we focus on the three summary metrics of the glucose dynamics.

Response half-lives ranged from 1h to 2.2h (Figure 3A) and average meal response heights ranged from 0.5 mmol/L to 1.5 mmol/L (Figure 3B). The posterior parameter distributions for each participant (Figure 3A-B) show the uncertainty associated with the parameter estimates for each participant; in some cases the distributions were overlapping between two individuals (and hence we lack sufficient resolution to claim inter-individual differences) while in other cases the distributions were clearly separated and would be significant within a Bayesian statistical testing framework (e.g. half-life comparing ID 20 and 23, Figure 3A) ^45^. Across participants we found a positive relationship between response half-lives and average meal heights, with slower glucose response half-lives associated with larger postprandial glucose spikes (R = 0.44, p = 0.02, Figure 3C). This suggested that postprandial glucose control (i.e. the height of glucose spikes following meals) depends on glucose clearance time, which might be determined physiologically by insulin sensitivity or beta cell function (Discussion).

**Figure 3.**
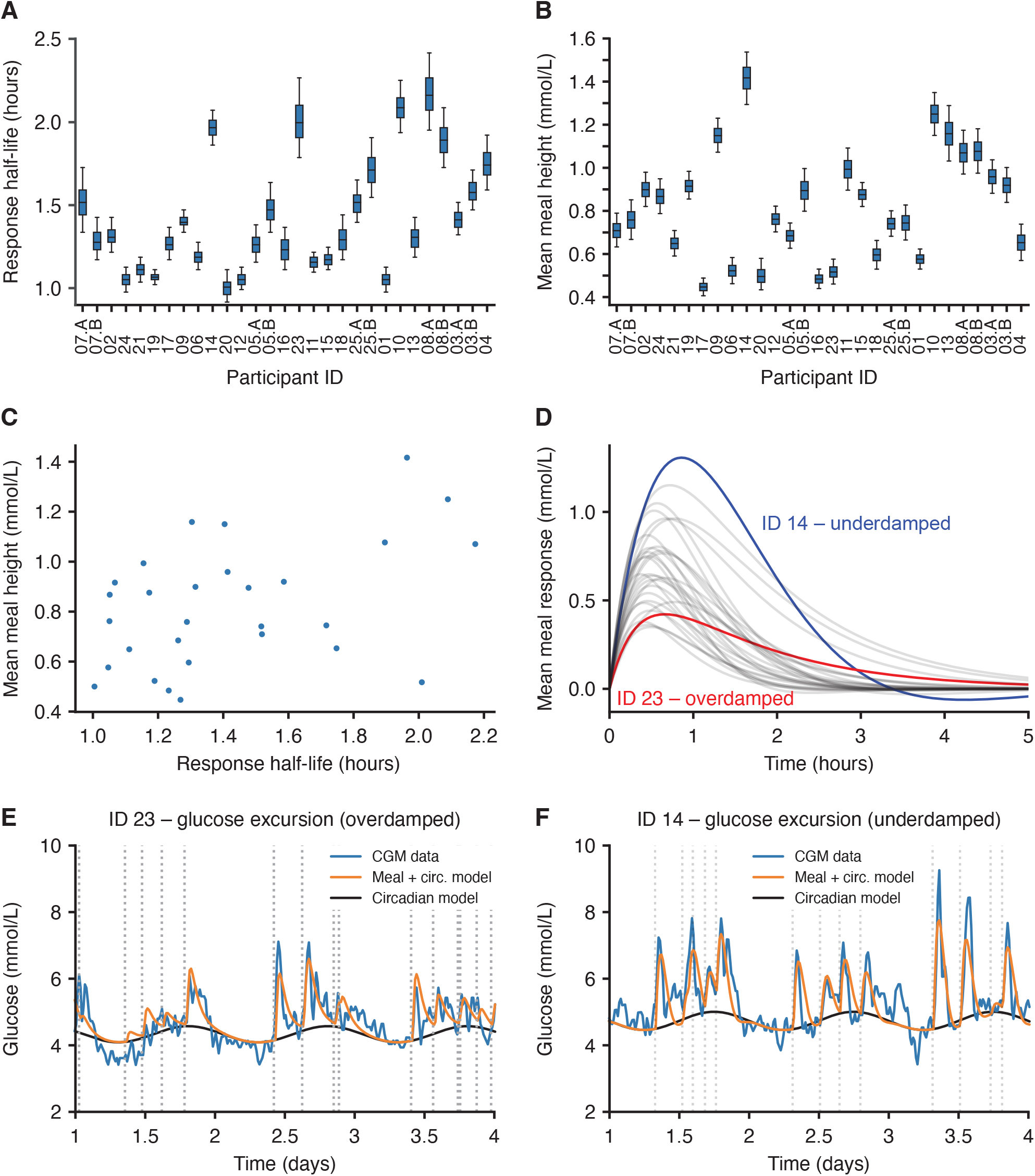
Characterising participant-specific post-meal glycaemic responses. (A) The inferred glucose response half-life for each participant, defined as the model-predicted time it would take for glucose levels to fall to 0.5 mmol/L following a peak of 1 mmol/L. The boxplots represent the 25th, median (50th) and 75th percentiles of the posterior distribution and the whiskers represent the 5th and 95th percentiles. (B) The average meal glucose spike height calculated as the mean height over all meals consumed during the experiment. (C) The average meal height as a function of the glucose meal response half-life. Points represent the mean posterior values for each participant. (D) Average meal response profiles using the posterior mean parameter values. (E-F) Examples comparing the CGM data (blue) with the model prediction incorporating circadian dynamics (black) plus meal consumption (orange) for two participants with overdamped and underdamped dynamics, respectively. The timestamps of meals are shown as dashed lines. Participant order is the same in Figures 3A-B and 4A-B.

The damping coefficients describing the shapes of glucose responses were clustered around 0 across all participants (Supplementary Figure 2), where values of 0 represent “critical” damping at the border between overdamped (non-dipping profiles, damping coefficient >0) and underdamped (profiles with a dip, damping coefficient <0). Interestingly, glucose responses were proposed to be critically damped in an early glucose model(Bolie, 1961), which would be consistent with our finding that the inferred values are scattered around zero. However, subject-to-subject variability is clearly found, with participant ID 14 showing a clear underdamped glucose response compared to the critically damped response in ID 23 (Figure 3D-F). The inferred meal response and circadian time functions (orange) are smoother than the glucose data (blue, Figure 3E-F), but the full model that also adds random fluctuations produces glucose traces that closely resemble the glucose data (Supplementary Figure 3).

The measured glucose coefficient of variation (CV), a metric of glycaemic control used in clinical settings^10,11^, showed significant associations with both the response half-lives (p=0.03) and average meal heights (p=0.01, R^2^ using both variables = 0.63). While the damping coefficient was not significantly associated with glucose CV, the individual shapes of glucose responses might play a role in other aspects of glucose dynamics such as overshooting and hunger^46^. Our results highlight that glucose response half-lives play a role in glycaemic control and may be a relevant metric for both fundamental research and clinical purposes.

### Circadian rhythms in baseline glucose levels show large inter-individual variability

In addition to the input from ingestion events, the model also allows for an underlying circadian rhythm in glucose levels described with three parameters: a baseline level that specifies the glucose at the trough of the oscillation, an amplitude parameter denoting the difference between the trough and peak of the oscillation, and the peak time of the oscillation.

The amplitudes of underlying circadian glucose rhythms were highly variable between participants (Figure 4A), being virtually null for some participants while exceeding 1 mmol/L for others (Figure 4C, ID 03 and 07, respectively). Notably, the parameter uncertainty was small enough that there was no overlap in the estimates for participants ID 03 and 07 (Figure 4A). To identify subjects whose profiles do not support a circadian baseline trend, we fitted an alternative model that lacked a circadian baseline and compared the two models using the Bayesian information criterion (BIC). For IDs 10, 13, 08, 25, 03 and 04 (which have the weakest amplitude according to Figure 4A), the BIC indicated evidence for the model lacking the circadian baseline, while the BIC favoured the model with an additional circadian component for all remaining participants (Supplementary Figure 4). The combination of the amplitude posterior estimates and heterogeneous model preference according to BIC suggests that circadian baseline glucose oscillations are highly unique physiological characteristics.

**Figure 4.**
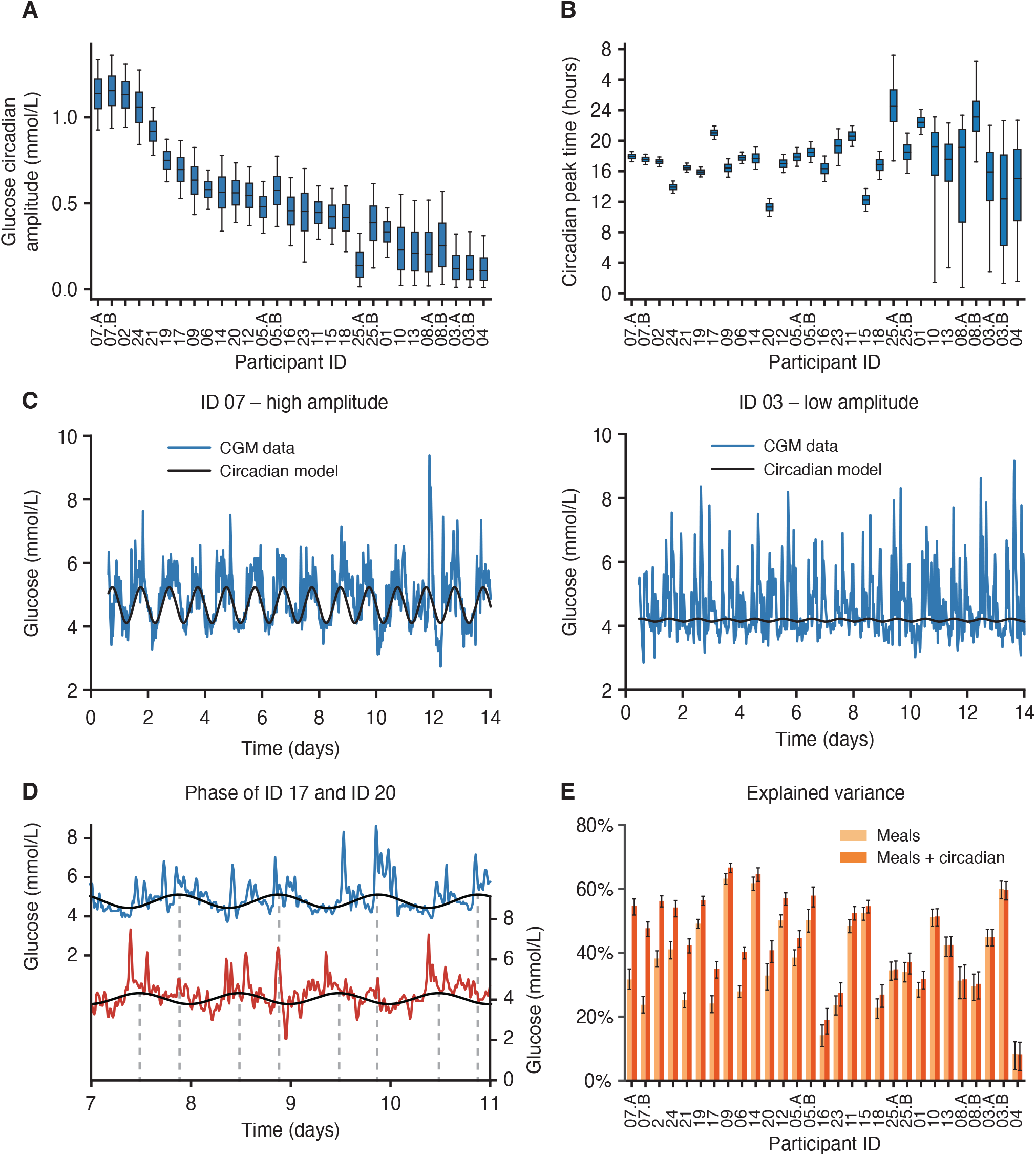
Amplitudes and peak times of circadian baseline levels of glucose are highly heterogeneous between participants. (A) The amplitude of the 24-hour sinusoidal circadian rhythm in baseline glucose levels after model fitting to the CGM data for all participants. The boxes represent the 25th, median (50th) and 75th percentiles of the posterior distribution and the whiskers represent the 5th and 95th percentiles. (B) The circadian phase of the glucose circadian rhythm across all participants. (C) Example of participants with a high (ID 07) and low (ID 03) amplitude glucose circadian rhythm. Blue, CGM data; black, fitted model of circadian baseline (using mean posterior parameter values). (D) Examples showing two participants with large phase difference in underlying glucose rhythm (ID 20 peak phase: 10:00, ID 17 peak phase: 20:00). (E) The explained variance in glucose levels using just the meal component of the model (light orange) compared with the inclusion of the circadian rhythm (dark orange). Error bars represent the 5^th^ and 95^th^ percentiles of the posterior distribution. Participant order is the same in Figures 3A-B and 4A-B.

The peak times of the glucose circadian oscillations similarly showed large differences between participants (Figure 4B, with the same participant order as in Figure 4A). While the peak times of the circadian underlying trend typically fell around the mid-afternoon, there were large phase differences between some individuals. For example, participant ID 20 had a peak time at 10:00 while it occurred much later for participant ID 17, falling at 20:00 (Figure 4D). The peak time distributions showed tight confidence intervals for participants with large amplitudes and wide intervals for participants with weaker amplitudes (Figure 4B). This relationship is probably caused by a low signal-to-noise ratio and suggests that the circadian peak time cannot be reliably inferred for participants with a low circadian amplitude.

Overall, the underlying circadian glucose rhythm can explain >15% of glycaemic variability in addition to the meal model for participants with large amplitudes (Figure 4E). While we have not tested whether it would be possible to modify either the peak time or amplitude of this rhythm, these personalised parameters should prove to be useful in applications such as personalised meal timing (Discussion).

### HR is well predicted by physical activity and circadian time but the predictability of HRV is highly variable between individuals

We next focused on the physical activity, HR and HRV data, where the aim was to model the dependencies between the variables and quantify the ability of a subset of the three signals to explain the variance of another, in addition to the contribution of circadian rhythms. For this, we created a new model (Model 2, Figure 2B) that incorporated the influence of physical activity on HR and HRV, and we used MCMC to sample from model parameters and quantify differences between individuals (all parameters shown in Supplementary Figure 5).

For HR the combination of circadian rhythms and physical activity as two inputs was consistently predictive, explaining 40-65% of HR variance across all participants (Figure 5A). Figure 5C-D shows an example of the predicted HR (orange) for two different participants using both the underlying circadian rhythm (black) and integrating the physical activity (green). While the circadian contribution to the explained HR variance differs for these two participants (Figure 5A), the correlation between the predicted and observed HR was ∼0.8 for both participants, demonstrating that time of day and activity state are necessary for optimal personalised modelling of HR, which is consistent with previous studies^47^.

**Figure 5.**
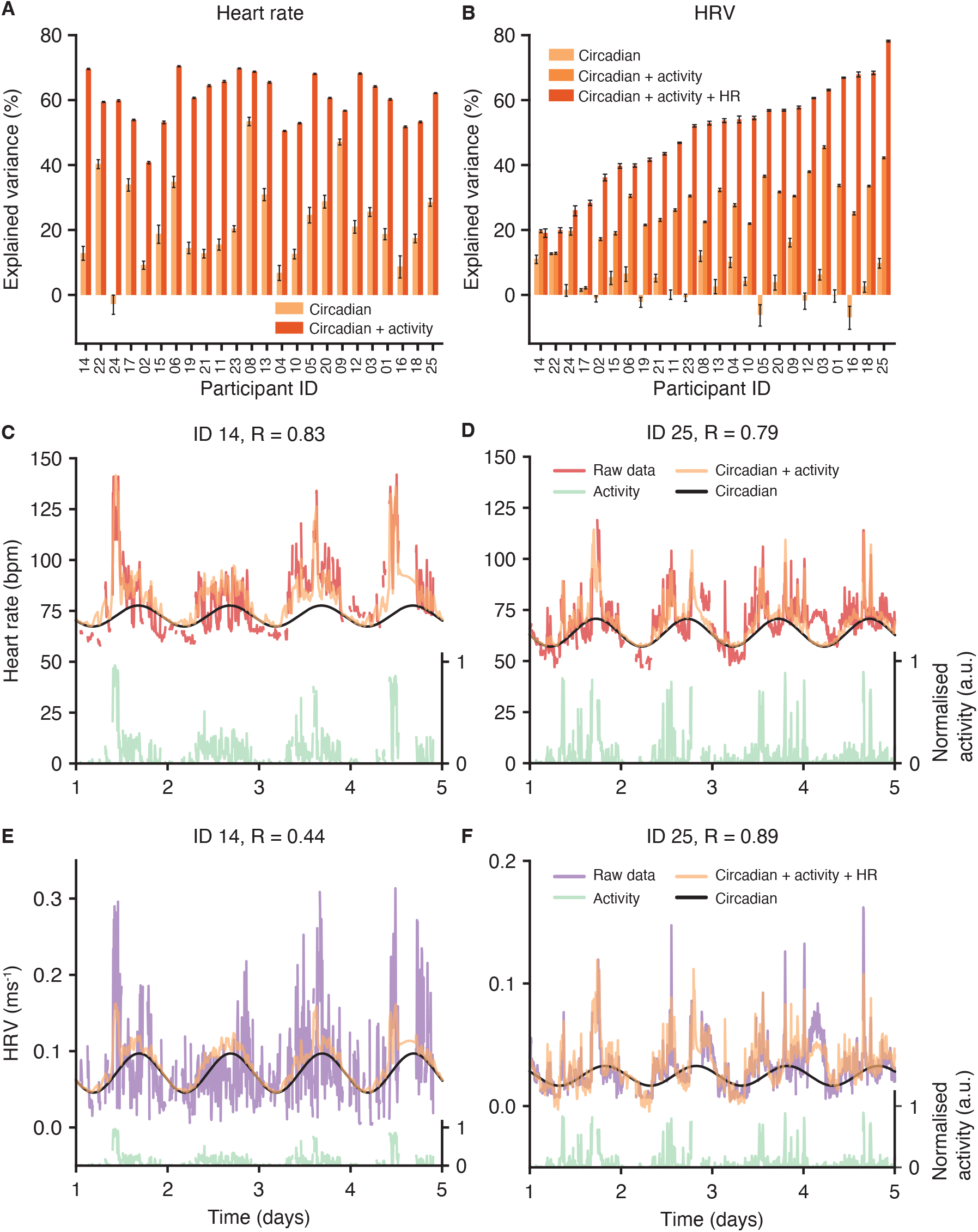
HRV predictions using multi-signal inputs and circadian rhythms are more heterogeneous than for HR. (A) The amount of variance of the HR signal explained by circadian rhythms (light orange) and a combined model with circadian rhythms and physical activity (dark orange). Error bars represent the 5^th^ and 95^th^ percentiles of the posterior distribution. (B) The amount of variance of the HRV (RMSSD^−1^) signal explained by circadian rhythms (light orange), a combined model with circadian rhythms and physical activity (medium orange) and a combined model with circadian rhythms, physical activity and HR (dark orange). (C,D) Examples comparing HR data with model predictions for two participants. Red: HR data; black: baseline circadian rhythm; green: physical activity (shown on normalised scale where 1 represents the maximum value); orange: model prediction with circadian rhythm and integrating activity. (E,F) Examples comparing HRV data with model predictions for two participants. Red: HRV data; black: baseline circadian rhythm; green: physical activity (shown on normalised scale where 1 represents the maximum value); orange: model prediction using circadian rhythms, physical activity and HR. Participant order is the same in panels A and B.

The ability to predict HRV was, in contrast, much more heterogeneous between participants with total variance explained between 20% and 80% (Figure 5B). This notable difference in predictability is illustrated with two participants, showing a favourable prediction for participant ID 25 (R = 0.89, Figure 2F) compared to ID 14 (R = 0.44, Figure 2E). In addition to inputs from physical activity and the underlying circadian rhythm, we evaluated whether the correlations between HR and HRV could be exploited by using HR to predict HRV (which is technically more difficult to measure than HR). The dependence between HR and HRV showed marked inter-individual differences, where for ID 25 the HR signal explained 40% of the variance compared to using just activity and circadian rhythm, but for ID 14 the addition of HR makes no difference to HRV prediction (Figure 5E-F). Given that HR and HRV receive different inputs from the SNS and PNS^25^, the strength of this dependence may be a function of the autonomic nervous system. Of note, ID 14 was previously diagnosed with diabetes currently treated only with lifestyle measures (and not pharmacological treatment) and autonomic dysfunction is a known complication of diabetes^48^.

### Integrating physical and heart activity signals helps explain glycaemic dynamics for some individuals

As a final modelling step, we integrated the physical and heart activity signals with the glucose-ingestion model to quantify how much of the glucose dynamics can be accounted for with physical activity, HR and HRV (Model 3, Figure 2C). To simplify the model inference problem, the parameters describing the physical and heart activity model in isolation (Model 2, Figure 2B) were locked to their posterior mean values, and we added three new parameters that describe the input of physical activity, HR and HRV on glucose levels, respectively (Model 3, Figure 2C). These influences were left unconstrained and could have a positive, negative or zero effect on glucose levels.

Fitting the parameters to the data revealed that the effect of physical activity on glucose (parameter *C*_5,1_) was generally negative, the effect of HR (parameter *C*_5,2_) was generally positive and the effect of HRV (parameter *C*_5,3_) was typically neutral across all participants (Figure 6A-C). Given that the raw data showed glucose spikes during some periods of exercise (Figure 1D), the negative influence of physical activity on glucose (*C*_5,1_) was not expected. To test the robustness of this prediction, we therefore refitted the data using three simpler models, where there was only one input at a time from the physical and heart activity signals (Supplementary Figure 6). The influence of physical activity on glucose remained negative even when it was the sole input from Model 2 into glucose levels, further suggesting that the overall effect of physical activity is to deplete glucose levels amongst the participants of our study. Meanwhile, as HR acts to increase glucose levels (*C*_5,2_), increased HR during intense exercise can still lead to a net increase in predicted glucose levels. Overall, the amount of additional explained variance contributed by the physical and heart activity signals was modest for most participants but contributed up to 15% in certain individuals (Figure 6D).

**Figure 6.**
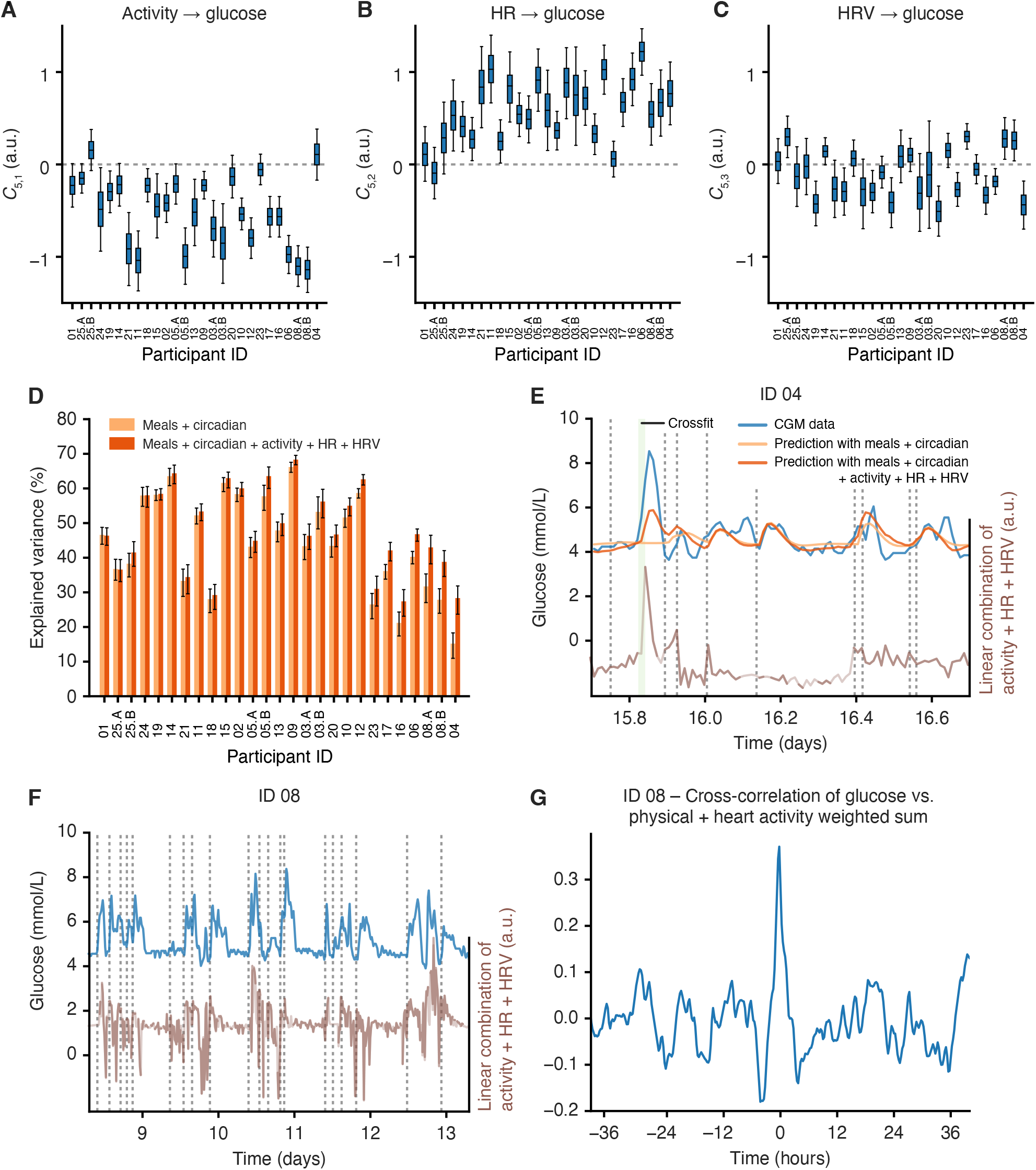
Adding physical activity, HR and HRV into the glucose dynamics model can help explain glucose dynamics. (A-C) Posterior distributions (shown as boxplots) of the model coefficients *C*_51_, *C*_52_, *C*_53_ across all participants, which correspond to the influence on glucose of physical activity, HR and HRV, respectively. The boxplots represent the 25th, median (50th) and 75th percentiles of the posterior distribution and the whiskers represent the 5th and 95th percentiles. (D) A comparison of the variance explained in the glucose signal using just the meal and circadian rhythm model (light orange) compared to the prediction that also incorporates physical activity, HR and HRV (dark orange). Error bars represent the 5^th^ and 95^th^ percentiles of the posterior distribution. (E) Example from ID 04 shows that the physical and heart activity data partially explain an exercise-induced glucose spike. Blue: glucose data; light orange: prediction using meal and circadian model components; dark orange: prediction including meal and circadian model components, physical activity, HR and HRV; green shade: recorded activity events; brown: the weighted sum of the physical activity, HR and HRV variables according to the inferred coefficients *C*_5,1_, *C*_5,2_ and *C*_5,3_; vertical dashed lines: ingestion events. (F) Example from ID 08 showing how glucose dynamics (blue) track with the weighted sum of the physical activity, HR and HRV variables according to the inferred coefficients *C*_51_, *C*_52_ and *C*_53_ (dark brown: data only; light brown: using filtered estimations from Model 2 to fill missing physical and heart activity data; vertical dashed lines: ingestion events). (G) the cross-correlation of the glucose (blue in (F)) with the weighted sum of the physical activity, HR and HRV variables (brown in (F)) using all recorded data. Participant order is the same between panels A, B and C.

Amongst the participants whose glucose dynamics benefit most from the physical and heart activity signals, the mode of action also differed. For participant ID 04, where the prediction of the glucose was the lowest with Model 1 (Figure 4E), the inclusion of the physical and heart activity signals allowed for partial prediction of exercise-induced glucose spikes that were otherwise absent from the glucose model that contained meals only (Figure 6E). For participant ID 08, which also showed a benefit of including physical and heart activity data, there were no notable isolated glucose spikes that were predicted by the physical and heart activity data (Figure 6F). Instead, the linear combination of the physical and heart activity variables (weighted according to the inferred coefficients *C*_5,1_-*C*_5,3_) appeared to track the glucose levels (Figure 6F), and the cross-correlation profile showed a maximum correlation between the two signals with no time delay (Figure 6G). This suggests that for this mode of action, the physical and heart activity signals make a more diffuse contribution to the prediction of glucose levels that is more related to baseline trends spread over the time series.

## Discussion

This study makes two major contributions to the quantitative analysis of glucose dynamics in terms of study design and computational analysis. Firstly, we used several wearable biosensors to measure multiple, interconnected data streams simultaneously. Without such high-resolution time series detailed dynamic inferences are impossible. On a practical level, one of the main advantages of health measurement with wearable biosensors is the rapidity of the experimental pipeline: in our study it took a team of two researchers two days to complete questionnaires and set up devices to record data for all 25 participants simultaneously. Secondly, we developed a data analysis method that combines stochastic dynamical modelling with Bayesian inference to learn personal parameters along with their associated uncertainty, and this parameter uncertainty was necessary to compare participants. The main insight of this personalised modelling is the clear inter-individual differences across a range of parameters and metrics, from the glucose half-lives and circadian oscillations, the coupling between HR and HRV, to the benefit of adding additional signals to predicting glucose.

The output of this modelling has wider implications for both understanding the biological underpinnings of cardiometabolic dysfunction as well as consequences for the use of wearables in a clinical setting. We first started by combining the food and drink events with the glucose time series, where we found that slow glucose dynamics are associated with large postprandial glucose spikes. Mechanistically, slow glucose disposal could relate to the quantity of ingested carbohydrates, the rate of gut absorption^49^, its metabolism^50^, suppression of endogenous glucose production^51^, insulin resistance or beta cell function^2^. Moreover, we also detected highly personalised circadian rhythms in the underlying glucose levels. While diurnal rhythms in beta cell function and insulin sensitivity have been shown at an average level within healthy populations^52–54^, it was unexpected to see such large differences in circadian glucose amplitude and phase between individuals. Future studies will determine whether this circadian glucose baseline trend is predictive of responses to specific meal times, e.g. in time-restricted eating, an intervention which restricts eating to a specific window within the 24-hour clock^55^.

Our results also have practical implications for clinicians as the physical and heart activity data explained up to 15% of glucose variability in our study, although this was highly variable between participants and may only be useful for some individuals. Never-the-less, from a clinical perspective, the incorporation of these additional signals might help both patients and clinicians understand glucose dynamics that seem otherwise disconnected from meal consumption (e.g. Figure 6E). As outlined in the Motivation paragraph, clinicians and patients with diabetes can link glucose excursion with ingestion events and intensive physical activity, but struggle to do this for the rest of the glucose dynamics measured over 24 hours.

There are multiple possible approaches to modelling our collected multi-modal data, and the particular glucose modelling methodology has often been historically dictated by the data available and by the stated goal^31–33^. Models based on differential equations range from simple, minimal models^34^ to more mechanistically detailed descriptions that include more variables, more spatial compartments and dozens of additional parameters^35^. Inspired by this work, there are many possible extensions that could be added to our glucose model, such as a meal-specific ingestion rates due to food content in carbohydrates, but also fat, fibre and protein content which are known to slow down nutrient absorption^14^, although the addition of meal-specific response shapes would effectively double the number of meal-related parameters.

An area that has seen a broad spectrum of time series models is in short-term glucose forecasting, typically for applications in closed-loop insulin delivery systems^42^. Gaussian state-space models that are conceptually similar to our own but with more variables have been deployed in artificial pancreas devices^56,57^, but a wide range of methods for short-range forecasting have also been used (reviewed in Woldaregay et al.^41^), including SVM and neural networks^36–38^. The advantage of such methods is that more complex nonlinear dependencies and long-range memory could possibly be captured. Here we traded some of this flexibility for explainability by using a relatively simple dynamical model with interpretable components and parameters, as our focus was on explaining the total time series rather than short-term forecasting. In the future, we envisage several applications, such as larger scale epidemiological studies (i.e. do inferred parameters track with health state?) and clinical trials to see whether parameters change in response to an intervention. Particularly for the second application, point estimation of parameters is not adequate, and uncertainty estimates are required to perform statistical tests for a given individual, which we achieve here through MCMC. In healthcare, there is increasing interest in the use of digital twins^58,59^ to integrate multiple clinical data streams, devise personalised treatments and perform risk modelling. As our approach contains interpretable parameters, it lends itself readily to exploring hypothetical situations by altering parameters (such as circadian amplitude or glucose response time) and simulating from the model. Overall, our method transforms a multivariable wearable data input into a series of metrics that describe the dependencies between physiological variables, including the relaxation timescales after external perturbations and circadian properties, and this approach provides a platform for probing physiological changes across circadian perturbations, ageing and cardiometabolic disorders.

### Limitations of the study

Our study population was young and in good health overall, and we lack additional, more detailed health information or standard clinical metrics such as HbA1c. We have therefore generally sought to identify differences between individuals without attempting to associate them with either good or bad health outcomes.

With respect to the modelling, a potential limitation of our approach is the use of a relatively simple linear model, which may not be able to capture more complex phenomena such as eventual decreases in hepatic glucose production during prolonged exercise^60^. A recent study based on deep learning found the addition of wristband activity data improved the root mean square error of 60-minute glucose forecasting by 2.25 mg/dL (0.1 mmol/L) from a baseline of 35.3 mg/dL (2.0 mmol/L), and hence more substantial improvements in glucose predictions may prove to be a difficult challenge even with more flexible models^61^.

## Methods

### Devices and experimental design

The Multi-Sensor Study (MSS) was approved by the local ethics committee (CER-VD, BASEC no. 2019-02245) and each participant signed a written informed consent. Recruitment was performed via posters at the École Polytechnique Fédérale de Lausanne (EPFL), Lausanne University Hospital (CHUV) and the University of Lausanne (UNIL) and via presentations given in the EPFL School of Life Sciences.

We included adults aged ≥ 18 years, with a smartphone compatible with the MyCircadianClock app (iOS or Android systems^43^) and able to take pictures of food/drinks, and who self-identified as disciplined enough and motivated to record all data for two weeks. The exclusion criteria were major illness/fever, surgery over the previous month, eating disorder, major mental illness, unable to give informed consent, taking medicines including paracetamol, aspirin or vitamin C supplements, enrolled in another interventional clinical trial (medication, medical device), shift work or travel to a different time zone before and during the study.

At baseline, we collected data on demographics, medical history, physical activity (short form of International Physical Activity Questionnaire, IPAQ-SF^62^, chronotype (The Munich ChronoType Questionnaire^63^, sleeping habits (Pittsburgh Sleep Quality Index^64^ and eating timing (with a custom questionnaire on eating habits during work and free days).

For each participant, we collected data for two weeks using the following devices: 1) Timestamps of food/drinks and text annotations collected with the smartphone application (app) MyCircadianClock^43^; 2) Continuous glucose monitoring (CGM) using the Abbott FreeStyle Libre Pro device; 3) Physical activity, heart rate (HR) and heart rate variability (HRV using RMSSD^−1^) using the CamNTech ActiHeart device version 5. Participants were instructed to take pictures of all consumed food and drink with the research-dedicated myCircadianClock smartphone. Recorded entries included a timestamped picture and a free-text annotation, and entries with the same annotation were considered as the same meal type. Participants could annotate photographs either immediately or in the following hours. Optionally, participants could type text-only entries without any picture, e.g., if the smartphone ran out of battery, or if it was not socially acceptable to take pictures in the current context. Participants were also asked to optionally log physical exercise using the app. While the CamNTech Actiheart device is waterproof, participants were permitted to briefly remove the device during showers and baths. Information on device technical failure, handling of missing data and data quality is included in the Supplementary Information.

### Pre-processing CGM data

We used nonparametric regression with Gaussian processes (GPs) to remove the long-term trends observed in the data. After mean-centring the data, we fitted a GP with a squared exponential kernel *K*_*SE*_(*t, t*′) = (−|*t* − *t*′|^2^/2*l*^2^) and a length scale *l*=48 hours using GPflow^65^.

### Data analysis

See the Supplementary Information for detailed computational methods which are summarised here. We use a linear Gaussian state space model (otherwise known as a Kalman filter^66^) to analyse the time series generated by the wearable devices, which was implemented using the ‘LinearGaussianStateSpaceModel’ distribution within TensorFlow Probability^67^. We will first describe the general data analysis framework before providing details on each of the three models used. For each model we define a dynamic model that describes the time evolution of the underlying physiological variables and a measurement model that incorporates measurement noise. For the dynamic model, we use a system of stochastic differential equations (SDEs)

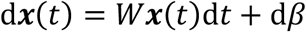

where *W* is a matrix describing the interactions between the variables *x*(*t*), and *β* is a brownian noise term with covariance matrix *Q*. The specific forms of *W* and *Q* are unique for each model and will be described below. To keep the model exact while benefiting from the generic framework of Gaussian state space models (a.k.a. as Kalman filters), we then convert this system of continuous-time SDEs into a model where time is discrete (see Supplementary Information for details).

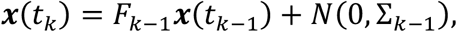

where *F*_*k*_ is the state-transition model and Σ_*k*_ is the covariance of the process noise. The measurement model describes the observation process and assumes that variables are observed with normally distributed measurement noise

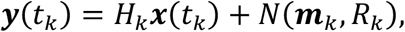

where *H*_*k*_ is the observation matrix and ***m***_***k***_ and *R*_*k*_ represents the mean and covariance of the observation noise, respectively. The goal is to use the wearable time series data ***y***_1:*T*_ to estimate parameters (denoted by ***θ***) for each participant. Within a Bayesian inference framework, the parameters of the model can be estimated from the data as follows

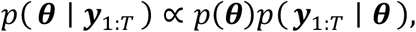

where *p*(***θ***) is the prior distribution of parameters and *p*(***y***_1:*T*_ | ***θ***) is the likelihood of observing the temporal data ***y***_1:*T*_ given the set of parameters ***θ***. Considering the time series sequence of data, the likelihood term for a given set of parameters ***θ*** can be expressed as

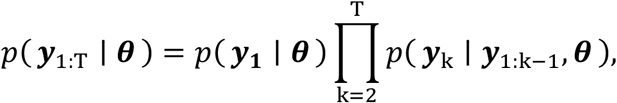

and the sequence of distributions *p*(***y***_**k**_ | ***y***_**1**:**k**−**1**_, ***θ***) are calculated within a Kalman filtering framework. Once the likelihood and priors are specified for each model, we used the Hamiltonian Monte Carlo sampler provided within TensorFlow Probability to sample model parameters from the posterior distribution using 4 different chains with 10,000 samples each. See Supplementary Information for further details on MCMC scheme. We then estimate the explained variance using 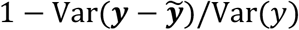, using the model predictions 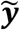 from the MCMC parameter samples.

### Model 1: glucose model

We model glucose dynamics (Figure 2A) with a two-dimensional system of SDEs, where the second variable *x*_GLUC2_ represents the glucose levels and the first variable *x*_GLUC1_ represents an unobserved latent variable that allows negative feedback within the system. In matrix form, the model is expressed as follows

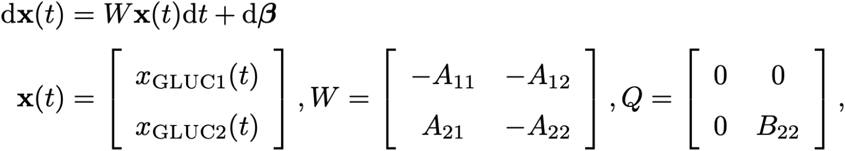

and where the coefficients *A*_*ij*_ are constrained to be positive. The covariance of the brownian noise term *β* is given by *Q*. The ‘damping coefficient’ is determined by whether the eigenvalues of the matrix *W* are real or complex. For the 2×2 matrix *W*, this damping coefficient can be determined by −det (*W* − *I* tr(*W*)/2)/(tr(*W*)/2)^2^.

We incorporate meal events (that are recorded at time *t*_*m*_) as producing a response function *r*_*m*_(*t*) by perturbing the first variable *x*_GLUC1_ to higher values (see Supplementary Information for precise functional form), and then the total meal function is the sum over all *M* individual meal responses

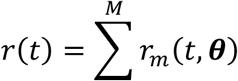

We define the glucose half-life parameter as the model-predicted time to return to 0.5 mol/L after a standardised increase of 1mmol/L. We also add an underlying circadian rhythm in glucose levels using a sinusoidal function

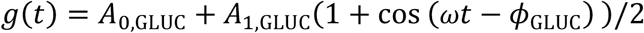

where *A*_0,GLUC_ is the baseline level, *A*_1,GLUC_ is the amplitude, *ω* is the frequency (fixed at 2*π*/24), and *ϕ*_GLUC_ is the peak time of the maximum. The observation model for the glucose model is then as follows

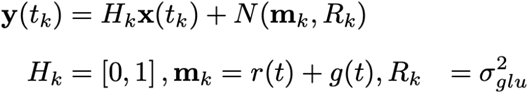

We compared Model 1 with an alternative version without circadian oscillations using the Bayesian Information Criterion (BIC) BIC = *k* ln(*n*) − 2 ln(*p*(***y***_1:*T*_ | ***θ***)), where *k* is the number of parameters and *n* is the number of data points. We calculated the difference in BIC score using Model 1 both with and without a circadian components and used a cut-off of ln 10 to indicate that the strength of evidence favoured a particular model^68^.

### Model 2 : physical and heart activity model

We model glucose dynamics (Figure 2B) with a with a three-dimensional system of SDEs, where the first variable *x*_ACT_ represents physical activity, the second variable *x*_HR_ represents heart rate and the third variable *x*_HRV_ represents heart rate variability, where we use the inverse of the root mean square of successive differences between normal heartbeats (RMSSD^-1^). We normalise all three variables by their respective standard deviations before inferring parameters. In matrix form, the model is expressed as follows

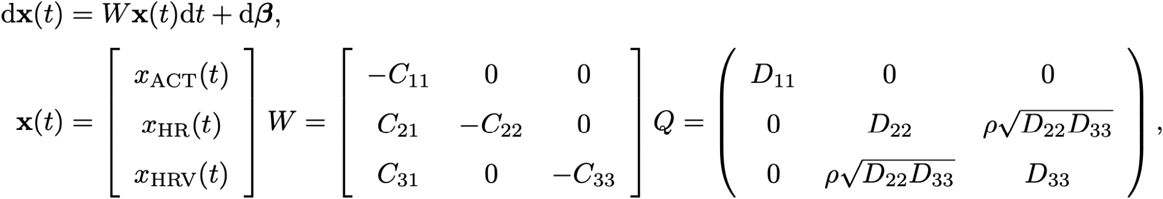

and where the coefficients *C*_67_ are constrained to be positive and the covariance of the brownian noise term *β* is given by *Q*. The observation model is then given by

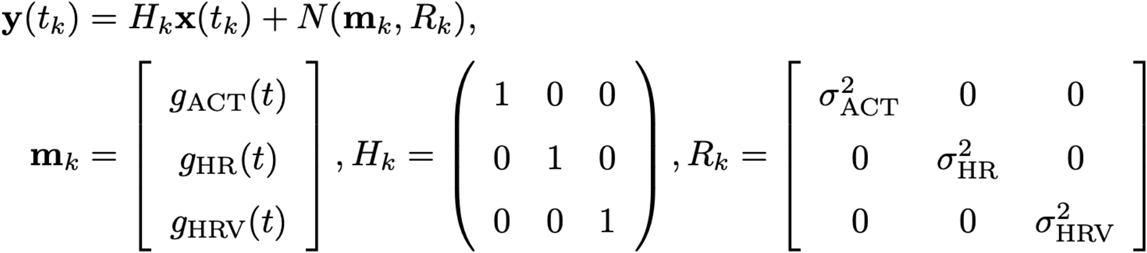

where *g*_ACT_(*t*), *g*_HR_(*t*) and *g*_HRV_(*t*) are circadian oscillatory functions (Supplementary Information).

### Model 3: combined model

The final model (Figure 2C) connects the CamNTech ActiHeart signals with CGM dynamics by stitching the previous glucose and physical and heart activity models together. Both models are otherwise left unchanged, but there is an introduction of three new parameters *C*_51_, *C*_52_ and *C*_53_ that describe the effect of physical activity, HR and HRV on glucose levels. These three parameters are left unconstrained and can take either positive or negative values.

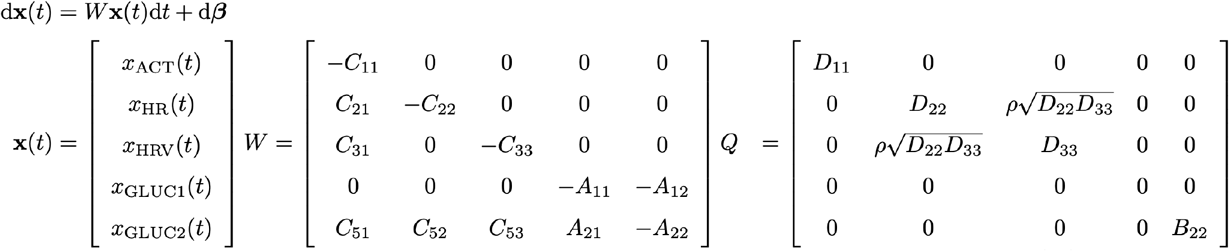

The observation model is then given by

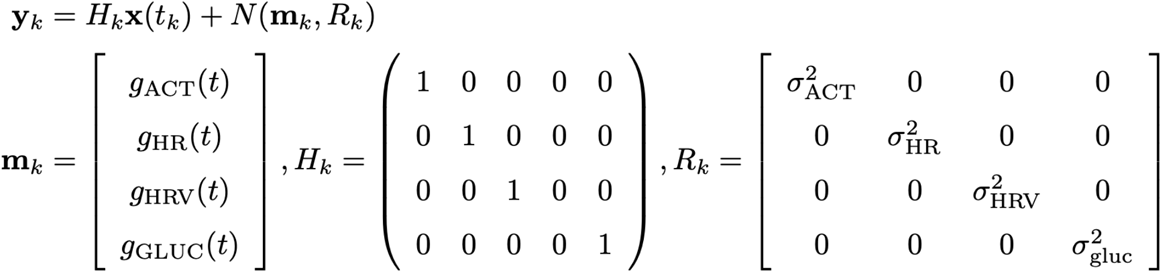

## Data Availability

The data will be made available after publication in an anonymised form.
The code for the method can be found at https://github.com/naef-lab/MultiSensor

## Acknowledgments

The authors wish to thank all participants in the study and the team of Professor Satchidananda Panda (Salk Institute) for the use of the *myCircadianClock* smartphone application. This project was supported by the grant #2018-427 of the Strategic Focal Area “Personalized Health and Related Technologies (PHRT)” of the ETH Domain to N.E.P. Dr. T-H Collet’s research is supported by grants from the Swiss National Science Foundation (PZ00P3-167826, 32003B-212559), the Leenaards Foundation, the Vontobel Foundation, the Nutrition 2000plus Foundation, the SwissLife Jubiläumsstiftung Foundation, the Swiss Society of Endocrinology and Diabetes, and the Swiss Multiple Sclerosis Society. The work in the Naef lab and purchase of the wearable devices was supported by the EPFL.

## Author contributions

Conceptualization: N.E.P., T.-H.C., F.N.; Methodology, N.E.P., T.-H.C., F.N.; Data collection: N.E.P., T.-H.C.; Computation and data analysis: N.E.P.; Data visualisation, N.E.P., T.H.C., F.N.; Writing: original draft preparation, N.E.P.; Writing: review and editing, N.E.P., T.-H.C., F.N.; Project administration: N.E.P., T.-H.C., F.N.; Supervision: T.-H.C., F.N.; Funding acquisition: N.E.P., T.-H.C., F.N. All authors have read and agreed to the published version of the manuscript.

## Data availability

The data will be made available after publication in an anonymised form.

## Code availability

The code for the method can be found at https://github.com/naef-lab/MultiSensor.

