## Supplementary Table, Methods and Figures for "Uncovering personalised glucose responses and circadian rhythms from multiple wearable biosensors with Bayesian dynamical modelling"

**3 References 16**

**4 Supplementary Figures 17**

### 1 Supplementary Tables

| Study ID | $MSF_{SC}$ (hours) | IPAQ (met-hours/week) | PSQI | App adherence (%) | Valid Actiheart |
|---|---|---|---|---|---|
| 01 | 4.8 | 13.65 | 7 | 100.0 | 72.5 |
| 02 | 5.5 | 9.65 | 2 | 100.0 | 58.6 |
| 03 | 3.3 | 5.30 | 1 | 100.0 | 59.7 |
| 04 | 4.9 | 15.30 | 7 | 100.0 | 80.5 |
| 05 | 5.0 | 37.65 | 3 | 100.0 | 85.5 |
| 06 | 5.3 | 9.49 | 4 | 100.0 | 73.3 |
| 07 | 3.3 | 35.30 | 4 | 100.0 | Not used |
| 08 | 4.2 | 24.95 | 5 | 100.0 | 70.3 |
| 09 | 3.7 | 2.47 | 7 | 100.0 | 93.7 |
| 10 | 4.6 | 53.10 | 7 | 100.0 | 90.8 |
| 11 | 3.6 | 3.10 | 3 | 100.0 | 72.4 |
| 12 | 5.6 | 28.95 | 2 | 100.0 | 94.4 |
| 13 | 4.5 | 14.47 | 9 | 100.0 | 83.4 |
| 14 | 2.3 | 10.95 | 2 | 100.0 | 74.3 |
| 15 | 4.7 | 23.30 | 5 | 100.0 | 41.4 |
| 16 | 3.6 | 39.30 | 5 | 100.0 | 89.6 |
| 17 | 4.5 | 5.00 | 5 | 83.3 | 82.6 |
| 18 | 5.0 | 25.10 | 4 | 100.0 | 85.6 |
| 19 | 3.8 | 18.60 | 4 | 100.0 | 91.3 |
| 20 | 4.2 | 9.60 | 8 | 100.0 | 89.2 |
| 21 | 4.5 | 14.16 | 5 | 100.0 | 60.9 |
| 22 | 5.8 | 3.22 | 6 | Not used | 81.3 |
| 23 | 4.3 | 24.95 | 7 | 100.0 | 86.3 |
| 24 | 2.5 | 28.82 | 5 | 100.0 | 74.6 |
| 25 | 4.2 | 9.65 | 5 | 100.0 | 91.8 |

**Table 1:** Chronotype and data quality metrics for study participants. $MSF_{SC}$ (hours): chronotype variable calculated from the Munich Chronotype Questionnaire (MCTQ); IPAQ: physical activity as measured with the International Physical Activity Questionnaire (IPAQ); PSQI: sleep quality as assessed with the Pittsburgh Sleep Quality Index. Range 0-27, where higher scores reflect lower quality sleep.; App adherence: the number of days with two recorded meals separated by at least 5 hours, calculated using all valid study days and excluding the first and last study days; Valid Actiheart (%): the % of Actiheart data that passes quality filtering for each participant.

### 2 Supplementary Methods

#### 2.1 Missing data and data quality

For each participant, we collected data for two weeks using the following devices: 1) timestamps of food/drinks and text annotations collected with the smartphone application (app) MyCircadianClock; 2) continuous glucose monitoring (CGM) using the FreeStyle Libre Pro device (Abbott); 3) physical activity, heart rate (HR) and heart rate variability (HRV using $\text{RMSSD}^{-1}$) using the Actiheart device (CamNTech). FreeStyle CGMs were replaced when devices fell off participants within the first week, which occurred 6 times, and data was not used when there was less than 48 hours of valid measurements. For one participant (ID 22), the FreeStyle CGM device failed without falling off, but due to device blinding this was not detected until the end of the experiment, and hence there is no glucose data for this participant. While the Actiheart device is waterproof, participants were permitted to briefly remove the device during showers and baths. At each time point the Actiheart produces an estimated quality of the signal (range 0-1), and we filtered the data based on a threshold of 0.8. After filtering the signal based on the quality, an average of 78.5% of the data was included, although this was specific to each participant (Supplementary Table 1). There was an Actiheart device failure for one participant (ID 07). The Actiheart data was exported with a time resolution of 5 minutes, while the FreeStyle CGM device records glucose every 15 minutes. To align the Actiheart and FreeStyle CGM time series, we chose the Actiheart measurement that was closest in time to the corresponding CGM measurement. This induces a maximum misalignment of 2.5 minutes, which we consider acceptable given that glucose typically has a response time >1 hour (Figure 3A).

#### 2.2 Modelling wearable time series data

We use a linear Gaussian state space model (otherwise known as a Kalman filter, see Särkkä and Solin (2019) for details) to analyse the time series generated by the wearable devices, and here we will first outline the general parameter inference framework before giving details for the specific models used. To account for both dynamics in underlying physiological variables changes as well as measurement error of the devices, this approach considers underlying physiological variables (referred to as latent state variables) $\mathbf{x}(t_k)$ (measured at times $t_k$) and observed variables $\mathbf{y}_k$. The linear Gaussian state space model of the wearable data has the following three components:

##### 2.2.1 Dynamic model

The dynamic model $\mathbf{x}(t_k) \sim p\left(\mathbf{x}(t_k) \mid \mathbf{x}(t_{k-1})\right)$ describes the transition probabilities of the underlying physiological variables. Our starting point is a continuous-time system of stochastic differential equations (SDEs)

$$\mathrm{d}\mathbf{x}(t) = W\mathbf{x}(t)\mathrm{d}t + \mathrm{d}\boldsymbol{\beta}, \tag{1}$$

where $W$ is a matrix describing the interactions between the variables and $\boldsymbol{\beta}$ is a brownian noise term with covariance matrix $Q$. The specific forms of $W$ and $Q$ are unique for each model and will be described below. We then convert this system of continuous-time SDEs into a model where time is discrete.

$$\mathbf{x}(t_k) = F_{k-1}\mathbf{x}(t_{k-1}) + N(0, \Sigma_{k-1}), \tag{2}$$

where $F_k$ is the state-transition model and $\Sigma_k$ is the covariance of the process noise. To perform this time discretisation we use the following two formulas, where details on their precise calculation for each model will also be specified below

$$\begin{aligned} F_k &= \exp\left(W\Delta t_k\right), \\ \Sigma_k &= \int_0^{\Delta t_k} \exp\left(W\left(\Delta t_k - \tau\right)\right) Q \exp\left(W\left(\Delta t_k - \tau\right)\right)^\top \mathrm{d}\tau, \end{aligned} \tag{3}$$

and where the time difference is defined as $\Delta t_k = t_{k+1} - t_k$.

##### 2.2.2 Initial distribution

The initial distribution $\mathbf{x}(t_0) \sim p\left(\mathbf{x}(t_0)\right)$ specifies the distribution of the physiological variables at the initial time $k = 0$. This is specified with $p\left(\mathbf{x}\left(t_0\right)\right) = N\left(\mathbf{x}\left(t_0\right) \mid \mathbf{m}_0, P_0\right)$, and where we use the stationary distribution of the SDE to define the mean and covariance $\mathbf{m}_0$ and $P_0$.

##### 2.2.3 Measurement model

The measurement model $\mathbf{y}_k \sim p\left(\mathbf{y}_k \mid \mathbf{x}(t_k)\right)$ describes the observation process, which again assumes a Gaussian distribution.

$$\mathbf{y}(t_k) = H_k\mathbf{x}(t_k) + N(\mathbf{m}_k, R_k), \tag{4}$$

where $H_k$ is the observation matrix and $\mathbf{m}_k$ and $R_k$ represents the mean and covariance of the observation noise, respectively.

#### 2.3 The marginal posterior distribution of parameters

The parameters of the model can be represented with the parameter vector $\boldsymbol{\theta}$, and the goal is to infer the parameters for each participant. Within a Bayesian framework, the parameters of the model can be estimated from the data as follows

$$p\left(\boldsymbol{\theta} \mid \mathbf{y}_{1:T}\right) \propto p(\boldsymbol{\theta})p\left(\mathbf{y}_{1:T} \mid \boldsymbol{\theta}\right), \tag{5}$$

where $p(\boldsymbol{\theta})$ is the prior distribution of parameters and $p\left(\mathbf{y}_{1:T} \mid \boldsymbol{\theta}\right)$ is the likelihood of observing the temporal data $\mathbf{y}_{1:T}$ given the set of parameters $\boldsymbol{\theta}$. Considering the time series sequence of data, the likelihood term for a given set of parameters $\boldsymbol{\theta}$ can be re-expressed as

$$p\left(\mathbf{y}_{1:T} \mid \boldsymbol{\theta}\right) = p\left(\mathbf{y}_1 \mid \boldsymbol{\theta}\right) \prod_{k=2}^{T} p\left(\mathbf{y}_k \mid \mathbf{y}_{1:k-1}, \boldsymbol{\theta}\right), \tag{6}$$

and the sequence of distributions $p\left(\mathbf{y}_k \mid \mathbf{y}_{1:k-1}, \boldsymbol{\theta}\right)$ are calculated within a Kalman filtering framework that we will now describe.

#### 2.4 Kalman Filter

We will now describe the Bayesian filtering approach used to estimate the likelihood term $p\left(\mathbf{y}_{1:T} \mid \boldsymbol{\theta}\right)$. Starting with the mean $\mathbf{m}_0$ and covariance $P_0$ of the initial distribution $p\left(\mathbf{x}\left(t_0\right)\right) = \mathrm{N}\left(\mathbf{x}\left(t_0\right) \mid \mathbf{m}_0, P_0\right)$, the Kalman filter recursion operates from the first to last data point $k = 1, 2, ..., T$. At each time point, there is first a prediction step that predicts the current state of the latent variables $\mathbf{x}\left(t_k\right)$ based on all previous measurements up to $\mathbf{y}_{k-1}$

$$p\left(\mathbf{x}\left(t_k\right) \mid \mathbf{y}_1, \ldots, \mathbf{y}_{k-1}\right) = \mathrm{N}\left(\mathbf{x}\left(t_k\right) \mid \mathbf{m}_k^-, P_k^-\right), \tag{7}$$

and where the predicted mean and covariance are calculated from

$$\begin{aligned} \mathbf{m}_k^- &= F_{k-1}\mathbf{m}_{k-1}, \\ P_k^- &= F_{k-1}P_{k-1}F_{k-1}^\top + \Sigma_{k-1}. \end{aligned} \tag{8}$$

The measurement $\mathbf{y}_k$ at time $t_k$ is then incorporated to create an updated estimation of the latent state $\mathbf{x}\left(t_k\right)$

$$p(\mathbf{x}(t_k) \mid \mathbf{y}_1, \ldots, \mathbf{y}_k) = \mathrm{N}(\mathbf{x}(t_k) \mid \mathbf{m}_k, P_k) \tag{9}$$

and where the updated mean and covariance are calculated using the update steps

$$\begin{aligned}
\mathbf{v}_k &= \mathbf{y}_k - H_k \mathbf{m}_k^-, \\
S_k &= H_k P_k^- H_k^\top + R_k, \\
K_k &= P_k^- H_k^\top S_k^{-1}, \\
\mathbf{m}_k &= \mathbf{m}_k^- + K_k \mathbf{v}_k, \\
P_k &= P_k^- - K_k S_k K_k^\top.
\end{aligned} \tag{10}$$

From these equations the terms necessary for the likelihood (Equation 6) can be calculated as a by-product

$$p(\mathbf{y}_k \mid \mathbf{y}_1, \ldots, \mathbf{y}_{k-1}) = \mathrm{N}\left(\mathbf{y}_k \mid H_k \mathbf{m}_k^-, S_k\right). \tag{11}$$

We will now specify the three models used, which allow calculation of the terms in Equation 11 and hence the likelihood using Equation 6. For each of the models we use the "LinearGaussianStateSpaceModel" distribution within Tensorflow Probability. This distribution requires the specification of the mean and covariance of both the dynamics and observation models, and the log likelihood is then calculated using the 'log_prob()' method.

#### 2.5 Model 1: glucose and meals model

We model glucose dynamics with a two-dimensional system of SDEs with two variables: $x_{\mathrm{GLUC1}}$ and $x_{\mathrm{GLUC2}}$. The second variable $x_{\mathrm{GLUC2}}$ acts as an input into the observation model to produce the measured variable $y$, which corresponds to the continuous glucose monitor (CGM) data. In matrix form, the model is expressed as follows

$$\begin{aligned}
&\mathrm{d}\mathbf{x}(t) = W\mathbf{x}(t)\mathrm{d}t + \mathrm{d}\boldsymbol{\beta} \\
&\mathbf{x}(t) = \begin{bmatrix} x_{\mathrm{GLUC1}}(t) \\ x_{\mathrm{GLUC2}}(t) \end{bmatrix}, W = \begin{bmatrix} -A_{11} & -A_{12} \\ A_{21} & -A_{22} \end{bmatrix}, Q = \begin{bmatrix} 0 & 0 \\ 0 & B_{22} \end{bmatrix},
\end{aligned} \tag{12}$$

where the coefficients $A_{ij}$ are constrained to be positive. Given the coefficients in the $W$ matrix, the positive constraints on the coefficients $A_{ij}$ ensures stability of the matrix (i.e. $-A_{11} - A_{22} < 0$ and $A_{11}A_{22} > -A_{12}A_{21}$). There are two principal reasons for modelling the glucose with two variables: 1) the inclusion of a second variable allows for negative feedback via the negative term $-A_{12}$, as negative feedback can arise due to the action of insulin; 2) when meals are consumed, we model this as impacting the first variable $x_{\mathrm{GLUC1}}$. As the first variable $x_{\mathrm{GLUC1}}$ acts to increase levels in the second variable $x_{\mathrm{GLUC2}}$ (via the parameter $A_{21}$), this leads to smooth, continuous increases in glucose after food consumption, which is a feature observed in the data. The covariance of the brownian noise term $\boldsymbol{\beta}$ is given by $Q$.

##### 2.5.1 State-transition model

The state-transition model $F_k$ can be calculated using a 2x2 matrix exponential

$$\begin{aligned} F_k &= \exp\left(W\Delta t_k\right), \\ &= e^{s\Delta t_k}\left(\left(\cosh(\sqrt{\gamma}\Delta t_k) - s\frac{\sinh(\sqrt{\gamma}\Delta t_k)}{\sqrt{\gamma}}\right)I + \frac{\sinh(\sqrt{\gamma}\Delta t_k)}{\sqrt{\gamma}}W\right), \end{aligned} \tag{13}$$

where

$$\begin{aligned} s &= (-A_{11} - A_{22})/2, \\ \gamma &= (-A_{11} + A_{22})^2/4 - A_{12}A_{21} = s^2 - \det(W). \end{aligned} \tag{14}$$

If $\gamma < 0$ then

$$\begin{aligned} \cosh\sqrt{\gamma}t &= \cos\sqrt{-\gamma}t, \\ \frac{\sinh\sqrt{\gamma}|t|}{\sqrt{\gamma}} &= \frac{\sin\sqrt{-\gamma}|t|}{\sqrt{-\gamma}}. \end{aligned}$$

##### 2.5.2 Covariance of the process noise

The covariance of the process noise $\Sigma_k$ can be calculated from the steady-state covariance $P_\infty$ as follows

$$\Sigma(\Delta t_k) = P_\infty - F_k(\Delta t_k)P_\infty F_k^\top(\Delta t_k), \tag{15}$$

where $P_\infty$ can be calculated symbolically using the following formula

$$WP_\infty + P_\infty W^\top + Q = \mathbf{0}. \tag{16}$$

##### 2.5.3 Incorporating meal events

The smartphone application provides a list of the recorded ingestion event times $\{t_m\}_{m=1}^{M}$ for a total of $M$ meals. We model the effect of meal event $m$ as causing an instantaneous increase of size $\delta_m$ to variable $x_{\text{GLUC1}}$. Given the delays from eating to reaching the insterstitial tissue measured by the CGM, we also include a time lag parameter $\tau$ for each participant, which is learnt in addition to the other parameters during model fitting. The response function $r_m(t, t_m, \boldsymbol{\theta})$ to a meal is then given by

$$r_m(t, t_m, \boldsymbol{\theta}) = \begin{cases} \Theta(t-\tau-t_m)\frac{A_{21}\,\delta_m\,\mathrm{e}^{s\,(t-\tau-t_m)}\sinh\left(\sqrt{\gamma}\,(t-\tau-t_m)\right)}{\sqrt{\gamma}}, & \gamma > 0 \\ \Theta(t-\tau-t_m)\frac{A_{21}\,\delta_m\,\mathrm{e}^{s\,(t-\tau-t_m)}\sin\left(\sqrt{-\gamma}\,(t-\tau-t_m)\right)}{\sqrt{-\gamma}}, & \gamma < 0 \end{cases}, \tag{17}$$

where the Heaviside function $\Theta(t-\tau-t_m)$ is defined as

$$\Theta(x) = \begin{cases} 1, & x > 0 \\ 0, & x \le 0 \end{cases}. \tag{18}$$

The meal function is then sum of the $M$ individual meals

$$r(t) = \sum^{M} r_m(t, t_m, \boldsymbol{\theta}). \tag{19}$$

To simplify the interpretation of the perturbation parameters $\delta_m$ (and to also make the definition of the prior simpler), we reparameterise the parameters $\delta_m$ to correspond to the maximum height of the glucose meal response. The time $t_{r_{\max}}$ of the maximum height of the meal response is

$$t_{r_{\max}} = \begin{cases} \tanh^{-1}(-\sqrt{\gamma}/s)/\sqrt{\gamma}, & \gamma > 0 \\ \tan^{-1}(-\sqrt{-\gamma}/s)/\sqrt{-\gamma}, & \gamma < 0 \end{cases}, \tag{20}$$

and the the maximum height of the glucose meal response is then given as

$$\max(r_m(t, t_m, \boldsymbol{\theta})) = \begin{cases} \frac{A_{21}\,\delta_m\,\mathrm{e}^{s\,(t_{r_{\max}})}\sinh\left(\sqrt{\gamma}\,(t_{r_{\max}})\right)}{\sqrt{\gamma}}, & \gamma > 0 \\ \frac{A_{21}\,\delta_m\,\mathrm{e}^{s\,(t_{r_{\max}})}\sin\left(\sqrt{-\gamma}\,(t_{r_{\max}})\right)}{\sqrt{-\gamma}}, & \gamma < 0 \end{cases} \tag{21}$$

A meal height parameter $\delta_M^*$ is then defined as $\delta_M^* = \max(r_m(t, t_m, \boldsymbol{\theta}))$, and the original perturbation parameter $\delta_m$ is then found by renormalising.

###### 2.5.4 Circadian dynamics

In addition to the effect of external meal events, we also model underlying circadian rhythms in glucose levels with a sinusoidal function

$$g(t) = A_{0,\mathrm{GLUC}} + A_{1,\mathrm{GLUC}}(1 + \cos(\omega t - \phi_{\mathrm{GLUC}}))/2, \tag{22}$$

where $A_{0,\mathrm{GLUC}}$ is the baseline level, $A_{1,\mathrm{GLUC}}$ is the amplitude, $\omega$ is the frequency (fixed at $2\pi/24$), and $\phi_{\mathrm{GLUC}}$ is the peak time of the maximum.

###### 2.5.5 Observation matrix

The observation matrix $H_k$ is such that only the second variable is observed

$$H_k = [0, 1]\,. \tag{23}$$

###### 2.5.6 Observation model

Finally, the observation model has the following form

$$\begin{aligned} \mathbf{y}_k &= H_k \mathbf{x}(t_k) + N(\mathbf{m}_k, R_k) \\ \mathbf{m}_k &= r(t) + g(t), R_k = \sigma^2_{\mathrm{GLUC}}. \end{aligned} \tag{24}$$

##### 2.5.7 Parameter description and priors

| Parameter | Description | Prior |
|---|---|---|
| $A_{0,\text{GLUC}}$ | Circadian baseline | $A_{0,\text{GLUC}} \sim$ half-normal(5) |
| $A_{1,\text{GLUC}}$ | Circadian amplitude | $A_{1,\text{GLUC}} \sim$ half-normal(1) |
| $\phi_{\text{GLUC}}$ | Circadian phase | $\phi_{\text{GLUC}} \sim \text{U}(0, 24)$ |
| $A_{1,1}$ | $x_{\text{GLUC1}}$ degradation rate | $\log(A_{1,1}) \sim \text{N}(0, 1)$ |
| $A_{1,2}$ | Suppression of $x_{\text{GLUC1}}$ by $x_{\text{GLUC2}}$ | $\log(A_{1,2}) \sim \text{N}(0, 1)$ |
| $A_{2,1}$ | Activation of $x_{\text{GLUC2}}$ by $x_{\text{GLUC1}}$ | $\log(A_{2,1}) \sim \text{N}(0, 1)$ |
| $A_{2,2}$ | $x_{\text{GLUC2}}$ degradation rate | $\log(A_{2,2}) \sim \text{N}(0, 1)$ |
| $\tau$ | Delay in meal response | $\tau \sim$ half-normal(0.5) |
| $B_{2,2}$ | Glucose diffusion noise | $B_{2,2} \sim$ half-normal(0.5) |
| $\delta_m^*$ | Meal-induced glucose increase | $\delta_m \sim$ half-normal(5) |

**Table 2:** Parameters Model 1: glucose and meals model.

#### 2.6 Model 2: physical and heart activity model

We model physical and heart activity dynamics with a three-dimensional system of SDEs, where the first variable $x_{\text{ACT}}$ represents physical activity, the second variable $x_{\text{HR}}$ represents heart rate and the third variable $x_{\text{HRV}}$ represents heart rate variability, where we use the inverse of the root mean square of successive differences between normal heartbeats (RMSSD$^{-1}$). We normalise all three variables by their respective standard deviations before inferring parameters. In matrix form, the model is expressed as follows

$$\begin{aligned} &\mathrm{d}\mathbf{x}(t) = W\mathbf{x}(t)\mathrm{d}t + \mathrm{d}\boldsymbol{\beta}, \\ &\mathbf{x}(t) = \begin{bmatrix} x_{\text{ACT}}(t) \\ x_{\text{HR}}(t) \\ x_{\text{HRV}}(t) \end{bmatrix} W = \begin{bmatrix} -C_{11} & 0 & 0 \\ C_{21} & -C_{22} & 0 \\ C_{31} & 0 & -C_{33} \end{bmatrix} Q = \begin{pmatrix} D_{11} & 0 & 0 \\ 0 & D_{22} & \rho\sqrt{D_{22}D_{33}} \\ 0 & \rho\sqrt{D_{22}D_{33}} & D_{33} \end{pmatrix}, \end{aligned} \tag{25}$$

where the coefficients $C_{ij}$ are constrained to be positive. The covariance of the brownian noise term $\boldsymbol{\beta}$ is given by $Q$. Dynamic changes and fluctuations in HR and HRV might be correlated, where this correlation is mediated by e.g. the parasympathetic nervous system. In the model, the correlation in the fluctuations between HR and HRV is quantified with the correlation parameter $\rho \in [-1, 1]$.

##### 2.6.1 State-transition model

The state-transition model $F_k$ can be calculated using a 3x3 matrix exponential

$$
\begin{aligned}
F_k &= \exp\left(W \Delta t_k\right), \\
&= \begin{pmatrix} \mathrm{e}^{-C_{11}\,\tau} & 0 & 0 \\ -\frac{C_{21}\,\mathrm{e}^{-C_{11}\,\tau}-C_{21}\,\mathrm{e}^{-C_{22}\,\tau}}{C_{11}-C_{22}} & \mathrm{e}^{-C_{22}\,\tau} & 0 \\ -\frac{C_{31}\,\mathrm{e}^{-C_{11}\,\tau}-C_{31}\,\mathrm{e}^{-C_{33}\,\tau}}{C_{11}-C_{33}} & 0 & \mathrm{e}^{-C_{33}\,\tau} \end{pmatrix}.
\end{aligned} \tag{26}
$$

###### 2.6.2 Covariance of the process noise

Similarly to the glucose model, the covariance of the process noise is calculated symbolically using the following formula

$$
W P_\infty + P_\infty W^\top + Q = \mathbf{0}. \tag{27}
$$

###### 2.6.3 Circadian dynamics

We model the underlying circadian rhythms in activity levels with a sinusoidal function

$$
g_{\mathrm{ACT}}(t) = C_{0,\mathrm{ACT}} + C_{1,\mathrm{ACT}}(1 + \cos(\omega t - \phi_{\mathrm{ACT}}))/2, \tag{28}
$$

where $C_0$ is the baseline level, $C_1$ is the amplitude, $\omega$ is the frequency (fixed at $2\pi/24$), and $\phi_{\mathrm{ACT}}$ is the phase of the maximum. Given that physical activity acts as an input into HR, the circadian function for HR receives an input from physical activity. We therefore model the circadian rhythms using two functions: a term that integrates input from physical activity $g^1_{\mathrm{HR}}(t)$ and an independent function $g^2_{\mathrm{HR}}$. The expression for the term $g^1_{\mathrm{HR}}(t)$ that integrates input from physical activity is then given by

$$
\begin{aligned}
\frac{d}{dt} g^1_{\mathrm{HR}}(t) &= C_{11} g_{\mathrm{ACT}}(t) - C_{22} g^1_{\mathrm{HR}}(t), \\
g^1_{\mathrm{HR}}(t) &= \frac{C_0\,C_{21}}{C_{22}} + \frac{C_1\,C_{21}}{2\,C_{22}} - \frac{C_1\,C_{21}\,\omega\,\sin\left(\varphi_{\mathrm{ACT}} - \omega\,t\right)}{2\,\left({C_{22}}^2 + \omega^2\right)} + \frac{C_1\,C_{21}\,C_{22}\,\cos\left(\varphi_{\mathrm{ACT}} - \omega\,t\right)}{2\,\left({C_{22}}^2 + \omega^2\right)}.
\end{aligned} \tag{29}
$$

We then model additional circadian component with a separate function

$$
g^2_{\mathrm{HR}}(t) = C_{0,\mathrm{HR}} + C_{1,\mathrm{HR}}(1 + \cos(\omega t - \phi_{\mathrm{HR}}))/2. \tag{30}
$$

The total circadian function is then a sum of these two components

$$
g_{\mathrm{HR}}(t) = g^1_{\mathrm{HR}}(t) + g^2_{\mathrm{HR}}(t). \tag{31}
$$

The same framework is then used for the HRV signal, with a term that integrates input from physical activity $g^1_{\mathrm{HRV}}(t)$ and an independent function $g^2_{\mathrm{HRV}}$. The expression for the term $g^1_{\mathrm{HRV}}(t)$ that integrates input from physical activity is then given by

$$\frac{d}{dt} g^1_{\mathrm{HRV}}(t) = C_{11} g_{\mathrm{ACT}}(t) - C_{22} g^1_{\mathrm{HRV}}(t)$$
$$g^1_{\mathrm{HRV}}(t) = \frac{C_0\, C_{31}}{C_{33}} + \frac{C_1\, C_{31}}{2\, C_{33}} - \frac{C_1\, C_{31}\, \omega\, \sin\left(\varphi_{\mathrm{ACT}} - \omega\, t\right)}{2\, \left({C_{33}}^2 + \omega^2\right)} + \frac{C_1\, C_{31}\, C_{33}\, \cos\left(\varphi_{\mathrm{ACT}} - \omega\, t\right)}{2\, \left({C_{33}}^2 + \omega^2\right)}. \tag{32}$$

We then model additional HRV circadian component with a separate function

$$g^2_{\mathrm{HR}}(t) = C_{0,\mathrm{HRV}} + C_{1,\mathrm{HRV}}(1 + \cos(\omega t - \phi_{\mathrm{HRV}}))/2. \tag{33}$$

The total circadian function is then a sum of these two components

$$g_{\mathrm{HRV}}(t) = g^1_{\mathrm{HRV}}(t) + g^2_{\mathrm{HRV}}(t). \tag{34}$$

###### 2.6.4 Observation matrix

The observation matrix $H_k$ is such that all three variables are observed

$$H_k = \begin{pmatrix} 1 & 0 & 0 \\ 0 & 1 & 0 \\ 0 & 0 & 1 \end{pmatrix}. \tag{35}$$

###### 2.6.5 Observation model

Finally, the observation model has the following form

$$\mathbf{y}(t_k) = H_k \mathbf{x}(t_k) + N(\mathbf{m}_k, R_k),$$
$$\mathbf{m}_k = \begin{bmatrix} g_{\mathrm{ACT}}(t) \\ g_{\mathrm{HR}}(t) \\ g_{\mathrm{HRV}}(t) \end{bmatrix}, R_k = \begin{bmatrix} \sigma^2_{\mathrm{ACT}} & 0 & 0 \\ 0 & \sigma^2_{\mathrm{HR}} & 0 \\ 0 & 0 & \sigma^2_{\mathrm{HRV}} \end{bmatrix}. \tag{36}$$

##### 2.6.6 Parameter description and priors

| Parameter | Description | Prior |
|---|---|---|
| $C_{0,\text{ACT}}, C_{0,\text{HR}}, C_{0,\text{HRV}}$ | Circadian baseline (activity, HR, HRV) | $C_{0,\text{ACT}}, C_{0,\text{HR}}, C_{0,\text{HRV}} \sim$ half-normal(1) |
| $C_{1,\text{ACT}}, C_{1,\text{HR}}, C_{1,\text{HRV}}$ | Circadian amplitude (activity, HR, HRV) | $C_{1,\text{ACT}}, C_{1,\text{HR}}, C_{1,\text{HRV}} \sim$ half-normal(1) |
| $\phi_{\text{ACT}}, \phi_{\text{HR}}, \phi_{\text{HRV}}$ | Circadian phase (activity, HR, HRV) | $\phi_{\text{ACT}}, \phi_{\text{HR}}, \phi_{\text{HRV}} \sim \text{U}(0, 24)$ |
| $C_{11}, C_{22}, C_{33}$ | Relaxation rate (activity, HR, HRV) | $C_{11}^{-1}, C_{22}^{-1}, C_{33}^{-1} \sim$ half-normal(0.01) |
| $C_{21}$ | Activation of $x_{\text{HR}}$ by $x_{\text{ACT}}$ | $\log(C_{21}) \sim \text{N}(0, 5)$ |
| $C_{31}$ | Activation of $x_{\text{HRV}}$ by $x_{\text{ACT}}$ | $\log(C_{31}) \sim \text{N}(0, 5)$ |
| $\rho$ | Correlation in diffusion between $x_{\text{HR}}$ and $x_{\text{HRV}}$ | $\rho \sim \text{U}(-1, 1)$ |
| $D_{11}, D_{22}, D_{33}$ | Diffusion noise (activity, HR, HRV) | $D_{11}, D_{22}, D_{33} \sim$ half-normal(5) |
| $\sigma_{\text{ACT}}, \sigma_{\text{HR}}, \sigma_{\text{HRV}}$ | Measurement noise (activity, HR, HRV) | $\sigma_{\text{ACT}}, \sigma_{\text{HR}}, \sigma_{\text{HRV}} \sim$ half-normal(1) |

**Table 3:** Parameters Model 2: physical and heart activity model.

#### 2.7 Model 3: combined model

The final model connects the physical and heart activity signals with glucose dynamics by connecting Model 1 (the glucose and meals model) with Model 2 (physical and heart activity). Three new parameters $C_{51}, C_{52}$ and $C_{53}$ are introduced that describe the effect of physical activity, HR and HRV on glucose levels ($x_{\text{GLUC2}}$), respectively. These three parameters are left unconstrained and can take either positive, negative or zero values.

$$\begin{aligned}
&\mathrm{d}\mathbf{x}(t) = W\mathbf{x}(t)\mathrm{d}t + \mathrm{d}\boldsymbol{\beta} \\
&\mathbf{x}(t) = \begin{bmatrix} x_{\text{ACT}}(t) \\ x_{\text{HR}}(t) \\ x_{\text{HRV}}(t) \\ x_{\text{GLUC1}}(t) \\ x_{\text{GLUC2}}(t) \end{bmatrix} W = \begin{bmatrix} -C_{11} & 0 & 0 & 0 & 0 \\ C_{21} & -C_{22} & 0 & 0 & 0 \\ C_{31} & 0 & -C_{33} & 0 & 0 \\ 0 & 0 & 0 & -A_{11} & -A_{12} \\ C_{51} & C_{52} & C_{53} & A_{21} & -A_{22} \end{bmatrix} \\
&Q = \begin{bmatrix} D_{11} & 0 & 0 & 0 & 0 \\ 0 & D_{22} & \rho\sqrt{D_{22}D_{33}} & 0 & 0 \\ 0 & \rho\sqrt{D_{22}D_{33}} & D_{33} & 0 & 0 \\ 0 & 0 & 0 & 0 & 0 \\ 0 & 0 & 0 & 0 & B_{22} \end{bmatrix}.
\end{aligned} \tag{37}$$

To simplify the model inference problem, the parameters from Model 2 describing the physical and heart activity model in isolation were locked to their posterior mean values. The covariance of the brownian noise term $\boldsymbol{\beta}$ is given by $Q$.

###### 2.7.1 State-transition model

Symbolic calculation of the matrix exponential required for the state-transition model becomes too cumbersome for Model 3, and we therefore use a Taylor series to approximate it (Moler and Van Loan, 2003).

$$\exp\left(W\Delta t_k\right) = \sum_{j=0}^{l} (W\Delta t_k)^j / j!, \tag{38}$$

along with the scaling property

$$e^{(W\Delta t_k)} = \left(e^{(W\Delta t_k)/m}\right)^m. \tag{39}$$

We use $l = 5$ and $m = 2$.

###### 2.7.2 Covariance of the process noise

For the covariance of the process noise, we use the Matrix Fraction Decomposition approach (see Särkkä 2006; Axelsson and Gustafsson, 2015) to solve

$$\frac{\mathrm{d}\Sigma(t)}{\mathrm{d}t} = W\Sigma(t) + \Sigma(t)W^\top + Q, \quad \Sigma(0) = \mathbf{0} \tag{40}$$

If we define matrices $C_\Sigma(\Delta t)$ and $D_\Sigma(\Delta t)$ such that $\Sigma(\Delta t) = C_\Sigma(\Delta t) D_\Sigma^{-1}(\Delta t)$ then we can now use matrix fractions to solve

$$\begin{aligned} \begin{pmatrix} C_\Sigma(\Delta t) \\ D_\Sigma(\Delta t) \end{pmatrix} &= \exp\left[\begin{pmatrix} W & Q \\ \mathbf{0} & -W^\top \end{pmatrix} \Delta t\right] \begin{pmatrix} \mathbf{0} \\ \mathbf{I} \end{pmatrix} \\ &= \begin{pmatrix} \exp(W\Delta t) & \Sigma(\Delta t)\exp(W\Delta t)^{-\top} \\ \mathbf{0} & \exp(W\Delta t)^{-\top} \end{pmatrix} \begin{pmatrix} \mathbf{0} \\ \mathbf{I} \end{pmatrix} \end{aligned} \tag{41}$$

As $D_\Sigma^{-1}(\Delta t) = \exp(W\Delta t)^\top$, there is no need for a matrix inversion.

###### 2.7.3 Observation matrix

The observation matrix $H_k$ is such that physical activity ($x_{\mathrm{ACT}}(t)$), HR ($x_{\mathrm{HR}}(t)$) and HRV ($x_{\mathrm{HRV}}(t)$) are observed along with the glucose levels ($x_{\mathrm{GLUC2}}(t)$).

$$H_k = \begin{pmatrix} 1 & 0 & 0 & 0 & 0 \\ 0 & 1 & 0 & 0 & 0 \\ 0 & 0 & 1 & 0 & 0 \\ 0 & 0 & 0 & 0 & 1 \end{pmatrix} \tag{42}$$

##### 2.7.4 Observation model

Finally, the observation model has the following form

$$\mathbf{y}_k = H_k \mathbf{x}(t_k) + N(\mathbf{m}_k, R_k)$$
$$\mathbf{m}_k = \begin{bmatrix} g_{\mathrm{ACT}}(t) \\ g_{\mathrm{HR}}(t) \\ g_{\mathrm{HRV}}(t) \\ g_{\mathrm{GLUC}}(t) \end{bmatrix}, R_k = \begin{bmatrix} \sigma^2_{\mathrm{ACT}} & 0 & 0 & 0 \\ 0 & \sigma^2_{\mathrm{HR}} & 0 & 0 \\ 0 & 0 & \sigma^2_{\mathrm{HRV}} & 0 \\ 0 & 0 & 0 & \sigma^2_{\mathrm{gluc}} \end{bmatrix} \tag{43}$$

##### 2.7.5 Parameter description and priors

Parameters for the physical and heart activity model (Model 2) are fixed at their posterior mean values, and the parameters for the glucose model (Model 1) are relearnt with the same priors. The priors for the 3 new parameters $(C_{51}, C_{52}, C_{53})$ in Model 3 are then as follows

| Parameter | Description | Prior |
|---|---|---|
| $C_{51}$ | Activation of $x_{\mathrm{GLUC2}}(t)$ by activity | $C_{51} \sim \mathrm{N}(0,1)$ |
| $C_{52}$ | Activation of $x_{\mathrm{GLUC2}}(t)$ by HR | $C_{52} \sim \mathrm{N}(0,1)$ |
| $C_{53}$ | Activation of $x_{\mathrm{GLUC2}}(t)$ by HRV | $C_{53} \sim \mathrm{N}(0,1)$ |

**Table 4:** Parameters Model 3: combined model.

#### 2.8 Posterior parameter sampling with MCMC

The parameter posterior distribution was sampled using Hamiltonian Markov Chain Monte Carlo (HMC), which uses the gradients of the posterior to improve the efficiency of the sampling. We used the 'HamiltonianMonteCarlo' function with TensorFlow Probability with 5 leapfrog steps, and we scaled the step size of each variable to approximately match the standard deviation of the posterior distribution. To achieve this, we sampled posterior parameters using two steps. Firstly, we sampled 10000 parameters (with a burn-in of 10000 samples) using the 'SimpleStepSizeAdaptation' kernel to select the global step size, which adapts the global step size to achieve a target acceptance probability of 0.75 (Andrieu and Thoms (2008)). We then scaled the the step size of each variable according to the standard deviation of this initial posterior distribution. Next, we resampled model parameters from the posterior distribution using 4 different chains with 10,000 samples each (with a burn-in of 10000 samples), again using 'SimpleStepSizeAdaptation' kernel to globally rescale the step size. The 'SimpleStepSizeAdaptation' kernel was only applied to first 80% of the burn-in samples.

### 4 Supplementary Figures

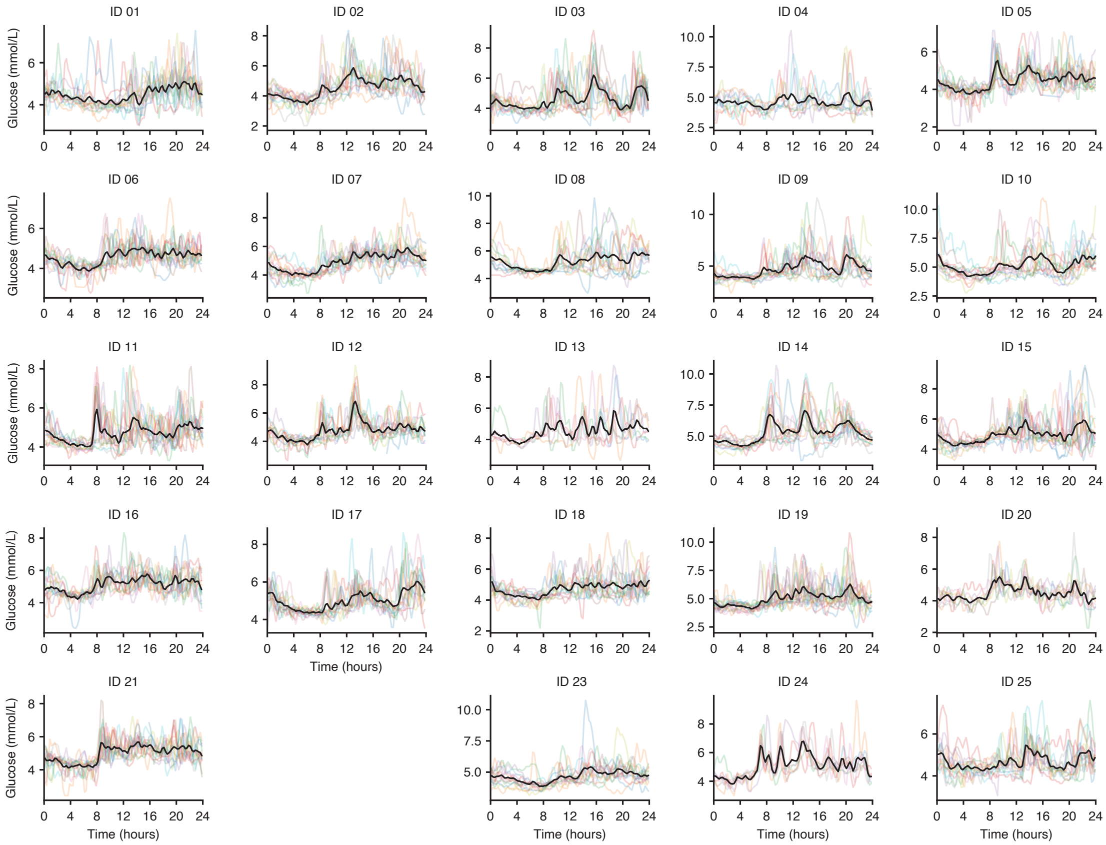

**Supplementary Fig. 1:** Glucose time-series data over a two-week period for all participants. Individual days are represented by different coloured lines and the mean glucose level over all measured days in black.

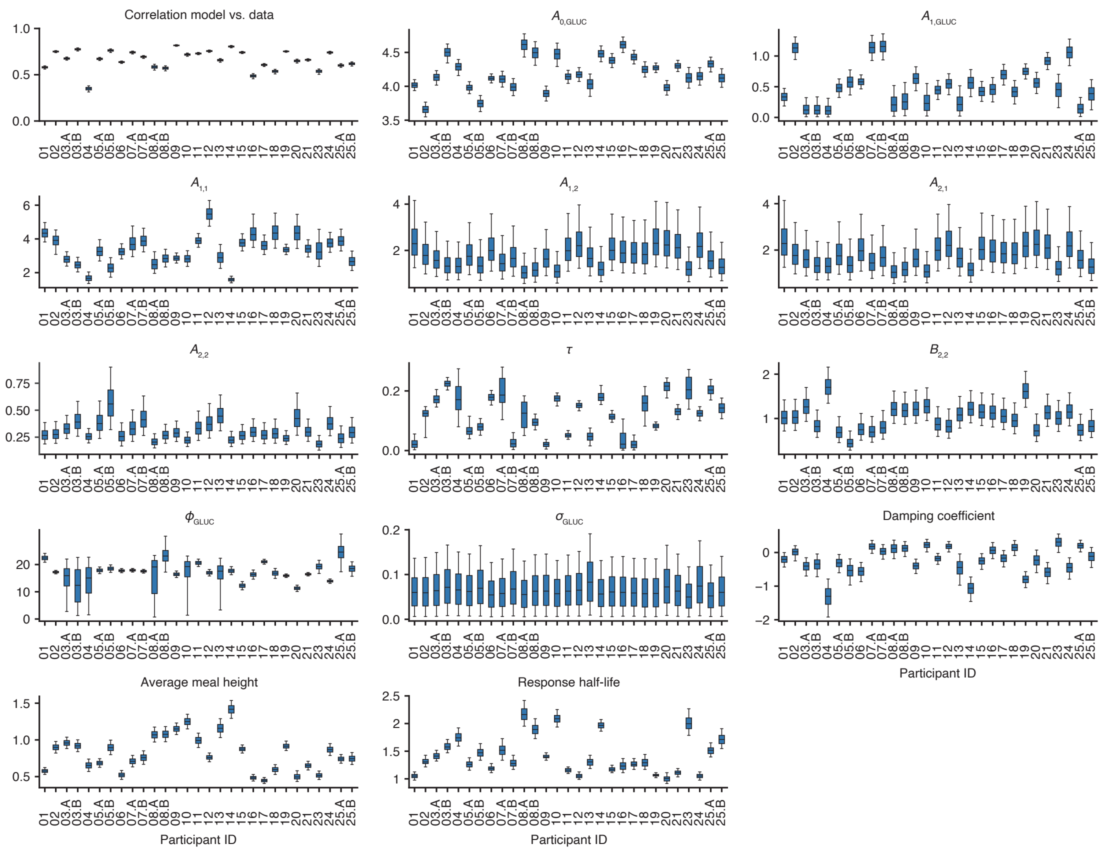

**Supplementary Fig. 2:** Posterior distributions (shown as boxplots) for all glucose model parameters (Model 1) across all participants. The first panel shows the correlation between the inferred model (including meals and circadian time) and the glucose CGM data for each participant. The boxplots represent the 25th, median (50th) and 75th percentiles of the posterior distribution and the whiskers represent the 5th and 95th percentiles.

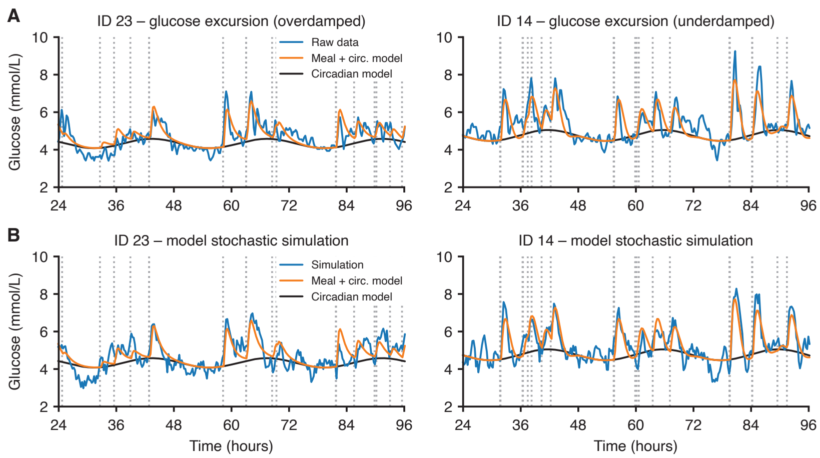

**Supplementary Fig. 3:** Simulating glucose traces using the full model creates realistic glucose traces. (A) Examples comparing the CGM data (blue) with the model prediction incorporating circadian dynamics (black) plus meal consumption (orange) for two participants with overdamped and underdamped dynamics, respectively. Same panels as Figure 3E-F, repeated here for easy comparison with panel B. (B) Simulations from the full glucose model that also includes random fluctuations (blue). The timestamps of meals are shown as black dashed lines.

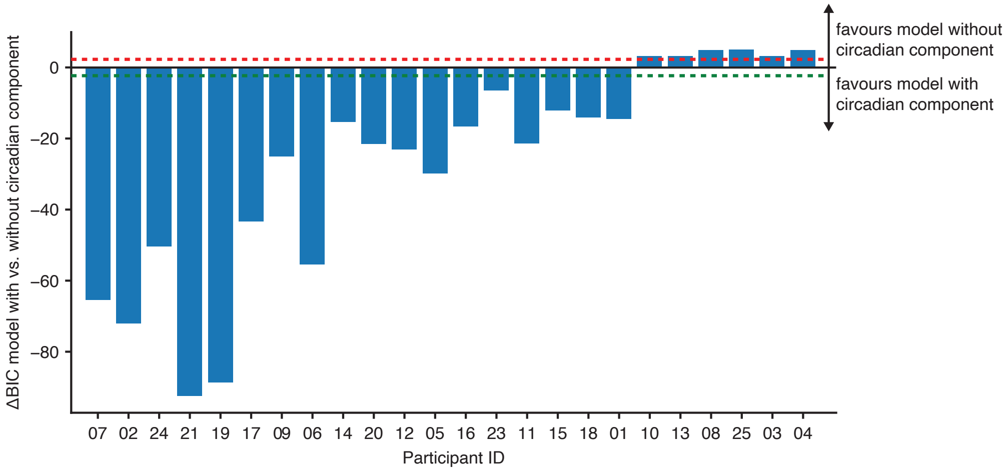

**Supplementary Fig. 4:** The difference in BIC score between Model 1 with a circadian component and Model 1 without a circadian component. The two models were first optimised to their maximum a posteriori probability (MAP) estimates, then the BIC score was calculated. A negative difference in BIC score favours the Model 1 with a circadian component, while a positive difference in BIC score favours the Model 1 without a circadian component. The dotted red line marks the threshold at which Model 1 without the circadian component is preferred, while the dotted green line marks the threshold at which Model 1 with the circadian component is preferred

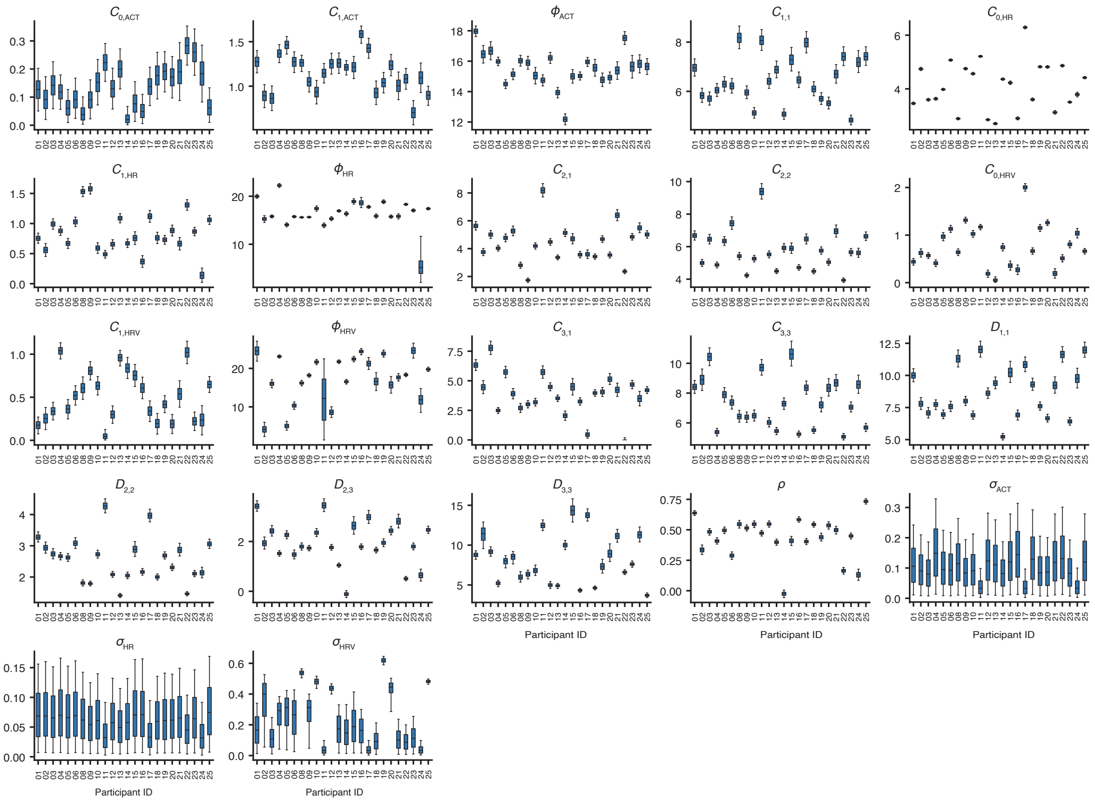

**Supplementary Fig. 5:** Posterior distributions (shown as boxplots) for all physical and heart activity model parameters (Model 2) across all participants. The boxplots represent the 25th, median (50th) and 75th percentiles of the posterior distribution and the whiskers represent the 5th and 95th percentiles.

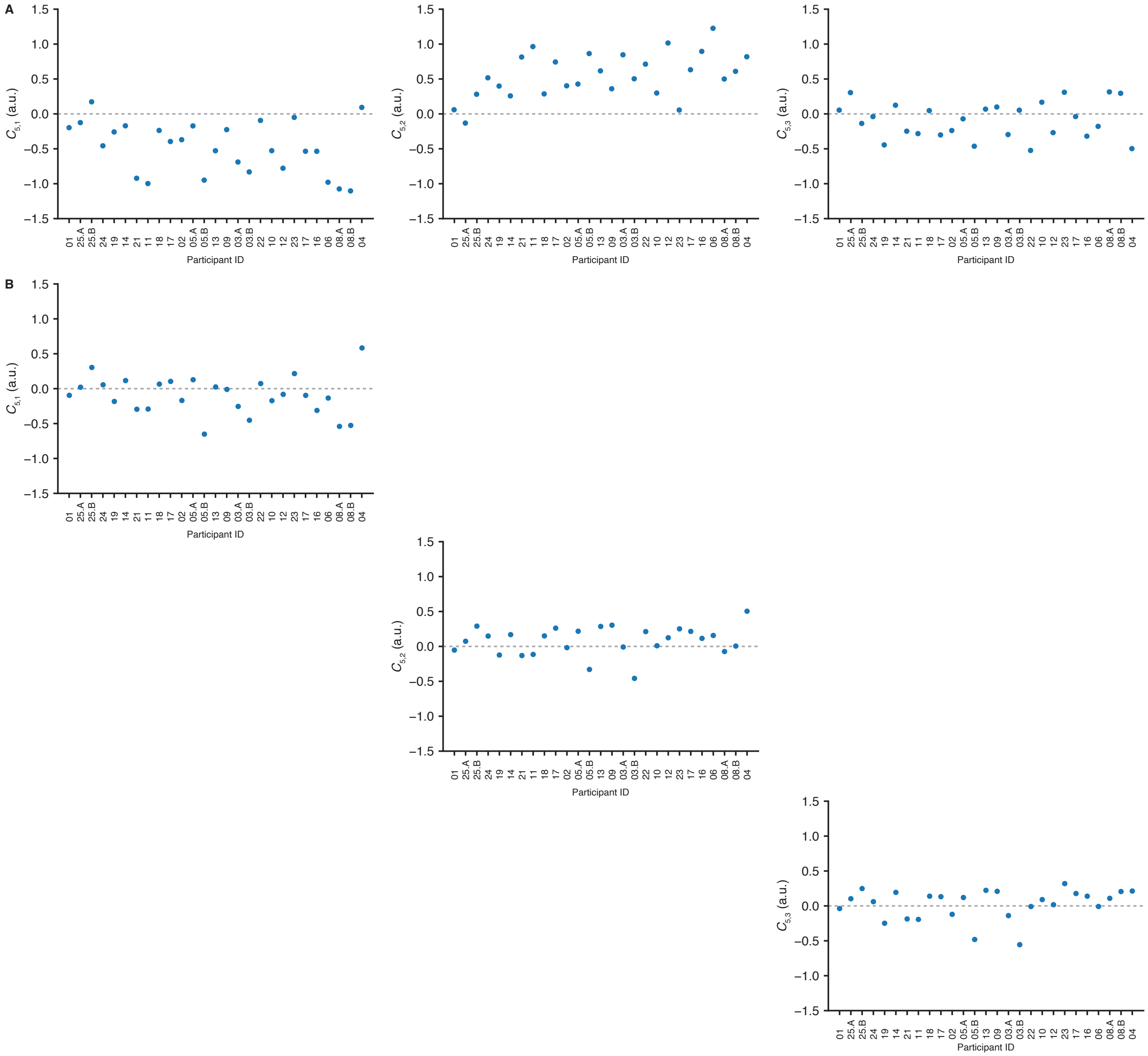

**Supplementary Fig. 6:** Model parameters when only one input from the physical and heart activity model is allowed. (A) Maximum a posteriori probability (MAP) estimate of coefficients $C_{51}, C_{51}$ and $C_{51}$ with all physical and heart activity inputs. (B) MAP estimate of the coefficients $C_{51}$ when physical activity (via parameter $C_{51}$) is the only physical and heart activity input into the glucose model. (C) MAP estimate of the coefficients $C_{52}$ when heart rate (via parameter $C_{52}$) is the only physical and heart activity input into the glucose model. (D) MAP estimate of the coefficients $C_{53}$ when heart rate variability (via parameter $C_{53}$) is the only physical and heart activity input into the glucose model.
